# Improving Polygenic Score Prediction for Underrepresented Groups Through Transfer Learning

**DOI:** 10.1101/2025.10.08.25337572

**Authors:** Hao Wu, Paulino Pérez-Rodríguez, Michael Boehnke, Yuehua Cui, Xiaoyu Liang, Ana I. Vazquez, Gustavo de los Campos

## Abstract

The advent of big data from GWAS consortia and biobanks led to remarkable improvements in polygenic score (PGS) prediction accuracy. However, most PGS were derived using data from Europeans (EU) and performed poorly when used to predict phenotypes of non-Europeans. Transfer Learning (TL) is a technique by which knowledge gained using data from one population is used to improve a model’s performance in another population. Here, we present GPTL, an R-package implementing three methods to build PGS using TL: gradient descent with early stopping, a penalized regression that shrinks variant effect estimates toward prior values, and a Bayesian model using a finite mixture prior that enables TL from multiple prior sources of information. Using simulated and real data from the UK-Biobank and All of Us, we showed that PGS derived using the TL algorithms implemented in the GPTL R-package performed better than PGS derived with EU or non-EU data only, and also outperformed (both in terms of accuracy and computational performance) commonly used methods to build PGSs using multi-ancestry data.

## Introduction

In the last two decades, genome-wide association studies (GWAS) have discovered hundreds of thousands of genetic variants associated with a wide spectrum of complex human traits and diseases. These findings inform the development of polygenic scores (PGS), which aggregate information across trait-or disease-associated genetic variants distributed across the genome. The advent of big data from GWAS consortia and biobanks has led to a remarkable increase in the number of trait-associated variants and a corresponding improvement in PGS prediction accuracy [1]. However, most PGS were derived using data from Europeans (EU) and have poorer performance when used to predict phenotypes of non-Europeans [2]. Despite recent investments in the collection of genomic data from non-Europeans, the underrepresentation of non-European ancestry in GWAS data will continue in the foreseeable future. Therefore, there is a need for methods that can leverage the large sample sizes available for Europeans to improve PGS prediction accuracy for non-Europeans.

Transfer Learning (TL) is a widely used machine learning technique to improve a model’s performance in a target population, leveraging knowledge gained from another population where the model has already performed well [3]. TL has been successfully applied to improve models for image-based diagnosis [4], drug sensitivity prediction [5], clinical trials [6], and multi-omics analysis [7, 8]. Importantly, TL algorithms enable improved performance of a model in one population by borrowing information from another population without requiring the sharing of individual-level data, which can be challenging and pose privacy concerns.

Recently, Zhao et al. [9] proposed using TL to improve PGS prediction accuracy for non-EUs. The TL algorithm used by Zhao et al. was gradient descent with early stopping (*TL-GDES*)–an approach commonly used in machine learning for TL and to prevent overfitting. *TL-GDES* uses variant effects estimated in one data set (typically a large data set of EU ancestry) as initial values to a gradient descent algorithm that, iterating on data from the target-ancestry group (e.g., African Americans), improves model fitness and PGS prediction accuracy for the target population. An early stopping rule is used to induce shrinkage of estimates towards the initial values, thus borrowing information between ancestry groups.

Penalized regressions and Bayesian methods are commonly used to induce the shrinkage of estimates. An extensive body of literature has documented important theoretical properties and good empirical performance of both approaches [10–12]. Commonly, Bayesian and penalized regression methods are formulated to shrink estimates towards null effects–this reduces the variance and (in many cases) the mean-squared error of estimates. However, simple modifications of these techniques can enable shrinkage towards prior estimates (e.g., EU-derived variant effects). Therefore, building on the well-established frameworks of penalized and Bayesian regressions, we developed two novel TL methods to develop PGS. The first method expands the traditional penalized regression framework by penalizing deviations of variant effect estimates from prior estimates (e.g., EU-derived variant effects). The second approach is a Bayesian model using a finite mixture prior that can be used for TL from multiple prior sources of information. We refer to these methods as Transfer Learning through Penalized Regression (*TL-PR*) and through Bayesian Mixture Model (*TL-BMM*).

To facilitate the construction of PGS using TL methods, we developed the GPTL (Genomic Prediction through TL, https://github.com/QuantGen/GPTL) R-package. The software offers functions that implement *TL-PR, TL-BMM*, and *TL-GDES* and can be used with individual genotype-phenotype data or summary statistics. The core of the algorithms is implemented in the C programming language [13] with a user-friendly R interface, which provides interoperability with other packages. The software scales well to PGS involving tens of thousands of variants when dense LD matrices are used and to millions of variants when the models are fitted using sparse LD reference panels and GWAS summary statistics.

In this study, after presenting the novel TL we developed, we benchmark the TL methods implemented in GPTL using data from African American (AA) and Hispanic ancestries in the All of Us study (AOU) [14] and EU ancestry data from the UK-Biobank [15] and AOU. Our simulations and real-data analysis results show that PGS derived using TL methods performed better than PGS derived using data from EU-ancestry or the target-ancestry (either AA or Hispanic) alone, and achieved a similar prediction performance as PGS derived using multi-ancestry data without the need to share individual genotype and phenotype data or large sufficient statistics. Furthermore, the three algorithms were computationally faster and achieved higher prediction accuracy than PRS-CSx [16] and PROSPER [17], which are widely used methods to develop PGS using multi-ancestry data.

## Results

A PGS is constructed by weighting allele counts or dosages at *p* variant with their association effect estimates, that is,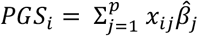 where x_*i* j_ ∈ {0, 1, 2} is the number of copies or dosage of a reference allele carried by individual *i* at variant *j*, and 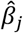 is the estimated effect of the reference allele at variant *j*. Multiple factors, notably including gene-gene interactions, genetic-by-environment interactions, and differences in allele frequencies and linkage disequilibrium (LD) patterns, can result in different variant effects across ancestry groups [2, 18, 19]. This can make the effect estimates derived using data from one ancestry group sub-optimal for the use of the PGS in another ancestry.

### Three Transfer Learning (TL) algorithms

Consider a simple setting where we have data from two populations: a target population (e.g., Hispanic) for which we wish to develop an accurate PGS, and a source population (e.g., EU-ancestry) from which we have a large data set that we wish to use to improve PGS performance in the target population. To allow for effect heterogeneity, we will use ***β***_s_ and ***β***_t_ to denote the (unknown) variant effects in the source- and target-ancestry groups, respectively. A TL algorithm derives estimates for the target-ancestry group 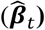 by shrinking those estimates towards estimates derived using data from the source-ancestry group 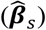.

#### Gradient descent with early stopping (TL-GDES)

In *TL-GDES* [9], variant effect estimates are derived by minimizing a loss function evaluated in data from the target-ancestry group. For a quantitative trait, the loss function can be a residual sum of squares

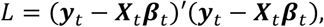

where ***β***_t_ is the unknown effect size in the target population, and ***y***_*t*_ and ***x***_*t*_ are the phenotype vector and the genotype matrix, respectively. A gradient descent algorithm produces a sequence of updates on regression coefficients, starting from initial values 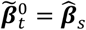 then in each cycle, each of the coefficients is updated, generating small changes that reduce the residual sum of squares. Specifically, updates are produced using 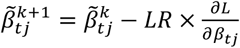 where 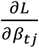 is the derivative of the loss function with respect to coefficient *j* evaluated in 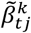 and LR is a learning rate parameter that controls the size of the updates. Because the objective function is strictly convex, when the algorithm is executed over enough cycles, the estimated effects converge to the solution of the optimization problem, in the above case, ordinary least squares (OLS) estimates. Stopping the algorithm before it converges to OLS estimates (aka early stopping) leads to estimates that are shrunk towards initial values 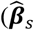 in the setting described above). The optimal number of cycles can be decided by monitoring PGS prediction accuracy over the iterations of the gradient descent algorithm in a calibration data set of the target-ancestry group. Further details about the algorithm are presented in [9] and in the Materials and Methods section.

#### Transfer Learning using penalized regressions (TL-PR)

Penalized regression methods are commonly used to prevent overfitting and to improve prediction performance [10]. In a standard penalized regression, coefficients are estimated by minimizing an objective function that includes a loss function (measuring the lack of fit of the model to the data) plus a penalty on model complexity. For a linear regression model, ***y***_*t*_ = ***x***_*t*_***β***_*t*_ + ***ε***, estimates are typically derived by minimizing with respect to *β*_t_ a penalized residual sum of squares of the form

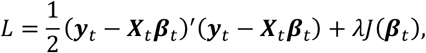

where λ (≥ 0) is a regularization parameter controlling the strength of the penalty, and J(***β***_t_) is a penalty function. Standard choices for the penalty function include the 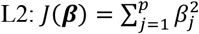 and 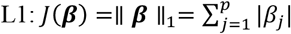 norms, and a linear combination of these norms, Elastic Net: 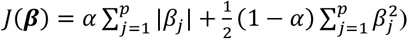. When these penalties are used, estimates are shrunk towards null effects.

To adapt penalized regression for TL, we propose using a novel penalty function that penalizes deviations from prior estimates, that 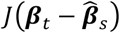. For the Elastic Net penalty, the objective function becomes

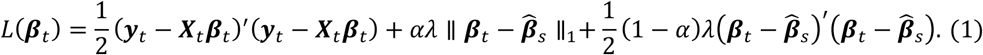

Estimates are then derived by minimizing L(***β***_t_) with respect to ***β***_t_.

For models using only an L2-penalty (α = 0), the value 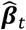 that minimizes the objective function of the *TL-PR* (1) can be shown to be

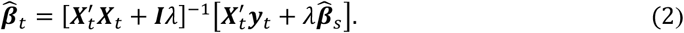

This estimator is similar to a Ridge regression estimator except that the estimates are shrunk toward prior values 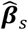 rather than zero.

When the L1-norm is involved in the penalty (0 < *α* ≤ 1), the minimizer of (1) does not have a closed-form solution. However, estimates can be obtained using a coordinate descent algorithm like the one used to solve the Elastic Net problem [11]. We derived the expressions needed to implement a coordinate descent algorithm for the *TL-PR* model and implemented them in the PR() function of the GPTL R-package. The resulting algorithm uses a soft-thresholding operator similar to the one used in a standard Elastic Net [11] with one key difference: instead of shrinking effects (completely or partially) towards zero, the update rules used in *TL-PR* shrink effects (also completely or partially) towards prior values. Further algorithm details are presented in the Supplementary Methods.

By design, *TL-GDES* and *TL-PR* transfer knowledge from a single source of prior information 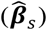. As more diverse sources of data become available, it seems reasonable to extend these approaches to allow TL from multiple sources of prior information 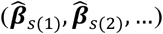 Previous studies [9, 23] have contemplated this situation by collapsing all the prior sources of information into a single vector of variant effects estimates (e.g., a weighted average, 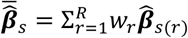 where 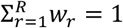 However, this approach assumes that the relative value of each source of information is the same across genomic regions. Therefore, to allow for a more flexible form of TL, we developed a Bayesian model using a finite mixture prior.

#### Bayesian model with an informative finite mixture prior (TL-BMM)

For most penalized regression methods, there is a Bayesian model whose posterior mode is equal to the solution of the corresponding penalized regression problem. For instance, the Bayesian estimator (2) is equal to the posterior mode of a Bayesian model with a Gaussian likelihood 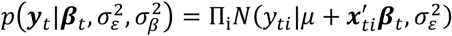 and a Gaussian prior for effects 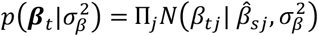 Here, N(⋅ | ⋅,⋅) stands for a normal density for the random variable at the left of the conditional bar (‘|’), and mean and variance are given by the expressions at the right of it.

To enable TL from multiple prior sources of information, we propose a Bayesian model using a finite mixture prior for variant effects. The prior includes one mixture component per source of information and potentially also includes one component to induce shrinkage toward zero. Specifically, assuming we have *R* sets of source estimates 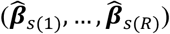 the finite mixture prior can be written as

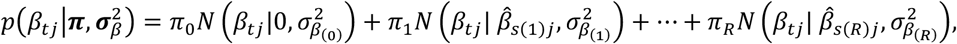

where *β*_*tj*_ is the effect of the variant *j* in the target population, *N*(⋅ | ⋅,⋅) denotes a normal density function, ***π*** = {*π*_0_, *π*_1_ …, *π*_*R*_} are prior mixture proportions satisfying 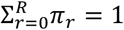 and 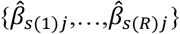 are the prior means for each of the components. To complete the specification of the mixture prior, we assigned a Dirichlet distribution to the prior mixture proportions (π) and scaled inverse-chi-squared priors to the variances of each of the components.

### The GPTL R-package

We implemented the three TL algorithms described above in the GPTL R-package. The core computations of each algorithm are implemented in the C programming language, with R functions used for the user interface. The functions GD(), PR(), and BMM() implement GDES, PR, and BMM, respectively. These functions take as input the sufficient statistics derived from genotype and phenotype data (***X***′***X, X***′***y***, the mean and variance of the phenotype, and the sample size used to compute the sufficient statistics). These sufficient statistics can be computed from individual genotype-phenotype data or from GWAS results and an LD reference panel. The function getSS() takes as inputs GWAS results and an LD reference panel and, after matching the variants in the LD reference panel with those in the GWAS table, it computes the sufficient statistics needed to run GD(), PR(), and BMM(). The matrix ***X***′***X*** can be dense (matrix class in R) or sparse (dgCMatrix class in package Matrix). Different C routines are internally dispatched for the dense and sparse cases.

The functions GD(), PR(), and BMM() return estimated effects or an entire path of values over gradient descent cycles in the case of GD() and over values of the regularization parameter in the case of PR(). Additional documentation and examples can be found in the GitHub repository of the package (https://github.com/QuantGen/GPTL). Toy examples demonstrating the use of each of the functions provided in the GPTL R-package are provided in the GitHub repository.

In the following sections, we present results from benchmarks where we compared the prediction performance of the TL methods implemented in the GPTL R-package with other methods. First, we present results from a pipeline using p-value thresholding (i.e., building PGSs using variants pre-selected based on GWAS results) involving both simulated and real data. For this pipeline, we present results from the three TL methods implemented in GPTL and three benchmarks (within-ancestry PGS, cross-ancestry PGS, and a PGS derived by combining data from multiple ancestries). Second, we present benchmarks using all the available variants and sparse LD reference panels. For this pipeline, we benchmark the methods implemented in the GPTL R-package against PROSPER and PRS-CSx.

### Benchmarks using variants selected through p-value thresholding

We used simulated and real data to benchmark the three TL methods. The target populations in which we evaluated prediction performance were African Americans (AA) and Hispanics from the All of Us (AOU) cohort. The source populations were from European ancestry, either European Americans (EA) from AOU, which we used in the simulations, or Europeans (EU) from the UK-Biobank, which were used in the real data analyses.

In this first benchmark, we compared the three TL methods implemented in the GPTL R-package against: (i) ***cross-ancestry* PGS prediction** which used EU-ancestry-derived PGSs to predict phenotypes of AA and Hispanics; (ii) ***within-ancestry* PGS prediction**, where we derived the PGSs using training data of the same ancestry as that of the prediction set; and (iii) ***combined-ancestry* PGS prediction** whereas the training data set included the data sets used to derive the *cross-ancestry* and *within-ancestry* PGSs. For this benchmark we used p-value thresholding (e.g., limited the number of variants used to those with a GWAS p-value < 1 × 10^−5^) and computed the sufficient statistics from individual genotype-phenotype data (another benchmark using GWAS results and an LD reference panel is presented later in this manuscript).

The pipeline used for this first benchmark consisted of three steps (**Figure 1**). In **Step 1**, we selected the variants to be used in the PGS using GWA analyses done with data from the source-ancestry (EU) and the training data from the target-ancestry group (either AA or Hispanic)– variants with GWA p-value < 1 × 10^−5^ were used to build PGS. Subsequently, in **Step 2**, we estimated variant effects using data from the source and target training data sets. Finally, in **Step 3**, we evaluated the prediction accuracy (the squared correlation between predictions and observations) in non-EU testing data that was not used in any of the preceding steps. Further details about each of these methods and the analysis pipeline are provided in the Materials and Methods section.

**Figure 1:**
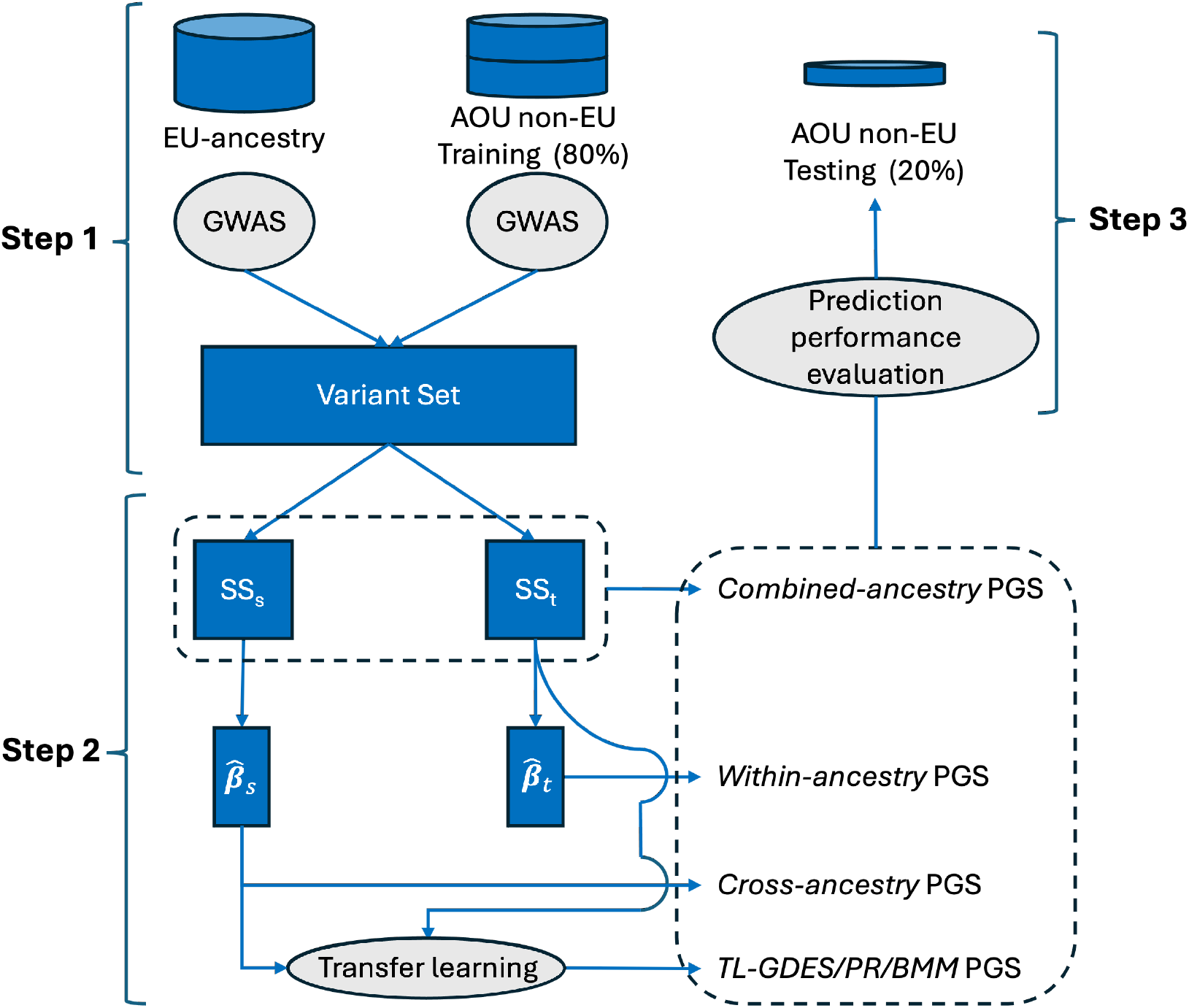
Graphical representation of the pipeline used to benchmark Transfer Learning (TL) algorithms for PGS derived using p-value thresholding. The pipeline used European (EU) and non-EU training data from the UK-biobank and the AOU cohorts to train six types of polygenic scores (*cross-ancestry, within-ancestry, combined-ancestry*, and three TL methods implemented in the GPTL R-package: *TL-GDES, TL-PR*, and *TL-BMM*). We evaluated the prediction accuracy in testing data from non-EU ancestry, either African Americans or Hispanics. The pipeline consists of three main steps: (1) variant selection based on GWAS results; (2) variant effect estimation using SS; and (3) evaluation of prediction accuracy in an independent testing data set. SS: Sufficient Statistics.

#### PGS prediction performance in simulations

The prediction performance of the six methods, *within-ancestry, cross-ancestry, combined-ancestry, TL-GDES, TL-PR*, and *TL-BMM*, varied between target-ancestry groups and simulation scenarios (**Figure 2**). In simulation settings where the correlation of causal effects between ancestries was very high (0.95, i.e., almost no effect heterogeneity at causal variants), the *cross-mancestry* PGS outperformed the *within-ancestry* prediction method except for AA when the training sample size for that ancestry group was large (N=50,000). This was as expected because of the larger sample size available from the EA cohort (N~151,000). On the other hand, when the correlation of causal effects was moderate (0.60), the *within*-*ancestry* prediction method outperformed the *cross-ancestry* prediction setting except for Hispanics when the sample size available from that ancestry group was small (N=10,000). In general, *cross-ancestry* prediction performed better in Hispanics than in AAs, which was expected since the portability of EU-derived effects is generally better in Hispanics than in AAs [19].

**Figure 2.**
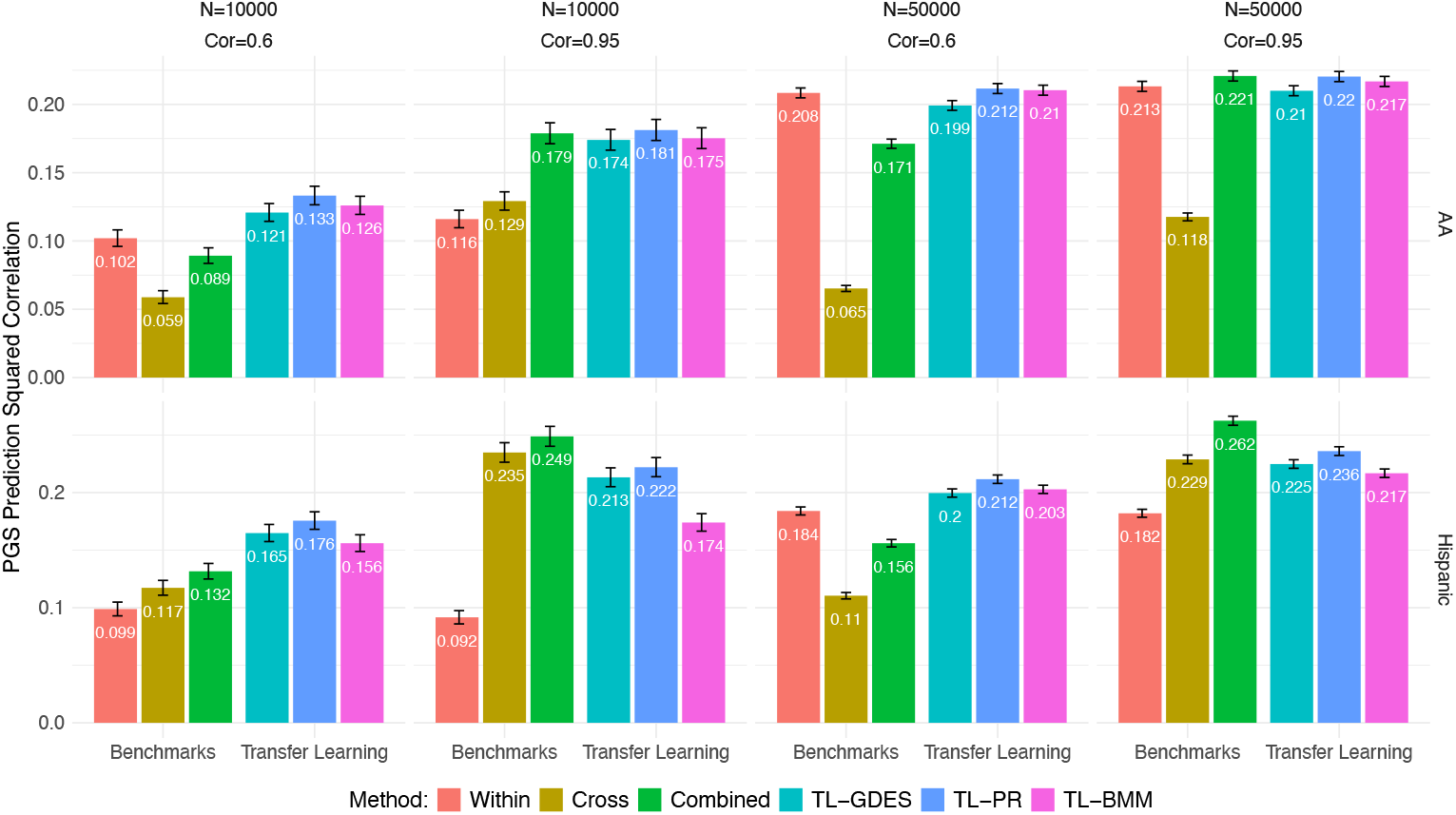
Prediction accuracy (± SE) of polygenic prediction methods in simulations, by method, sample size of the non-European training set (N), correlation of causal effects (Cor), and the ancestry of the testing data set. *Within* uses training data from AOU from the same ancestry as the testing data set. *Cross* uses data of European American (EA) ancestry. *Combined* uses the data used in *Within* and *Cross*. The three Transfer Learning (TL-) methods use the estimates obtained with EA data (*Cross*) as priors to a TL algorithm: Gradient Descent with Early Stopping, *GDES*, Penalized Regression, *PR*, or Bayesian Model, *BMM*, applied to the data set used in the *Within* method.

Across scenarios, the *combined-ancestry* analyses, which use a large sample size of EA ancestry as well as data from the target-ancestry group, tended to outperform both the *within-* and *cross-ancestry* PGSs. The only exception was the scenario with a very low correlation of effects and a large training sample size (N=50,000) from the non-EA ancestry group; in these cases, the *within-ancestry* PGS outperformed both the *cross-ancestry* and *combined-ancestry* methods.

Across scenarios, the TL methods had similar or, in many cases, slightly better prediction performance than the *combined-ancestry* prediction. When the correlation of causal effects was high (0.95), TL methods performed similarly to the *combined-ancestry* PGS. However, when the correlation of causal effects was moderate (0.60), TL methods outperformed the *combined-ancestry* PGS. Among the three TL methods, the Penalized Regression was the best-performing TL method, followed by the Bayesian Mixture Model, and then the Gradient Descent with Early Stopping. As one would expect, increasing the sample size of the non-EA led to an improvement in prediction for the methods that used those data (*within-ancestry, combined-ancestry*, and TL methods). The impact of sample size on prediction accuracy was particularly clear for AA (Figure 2).

#### PGS prediction performance with real data

When using real phenotypes, the *cross-ancestry* PGS predictions were typically better than *within-ancestry* PGS (see **Table 1** for results averaged across 11 traits, and **Figure 3** and **Table S1** for trait-specific results). This was expected given the much larger sample size of the UK-Biobank data. However, the *combined-ancestry* PGSs had better prediction accuracy than *cross-* and *within-ancestry* PGSs. Averaged across the eleven traits that we evaluated, the UK-Biobank-derived PGS (*cross*-*ancestry*) had a squared correlation 137% higher than the *within*-*ancestry* PGS in AA (124% higher in Hispanic), and the one that used both sources of data (*combined-ancestry*) had a squared correlation 235% higher than the *within*-*ancestry* in AA (197% higher in Hispanic) (Table 1). The three TL methods outperformed the *within*- and *cross-ancestry* PGSs by a sizable margin (~210% gain in AA and ~185% in Hispanics), and achieved very similar prediction squared correlation as the *combined-ancestry* PGS (Table 1).

**Table 1.** Average squared correlation (across eleven real phenotypes) by the ancestry of the testing set and the method used.

| Target-Ancestry Group | Method | Average squared correlation (SE) | % Change Relative to |  |  |
| --- | --- | --- | --- | --- | --- |
|  |  |  | *Within-ancestry* | *Cross-ancestry* | *Combined-ancestry* |
| African American | *Within-ancestry* | 0.008 (0.001) | --- | -58% | -70% |
|  | *Cross-ancestry* | 0.019 (0.002) | 137% | --- | -29% |
|  | *Combined-ancestry* | 0.027 (0.002) | 235% | 41% | --- |
|  | *TL-GDES* | 0.025 (0.002) | 205% | 29% | -9% |
|  | *TL-PR* | 0.026 (0.002) | 216% | 33% | -6% |
|  | *TL-BMM* | 0.026 (0.002) | 223% | 36% | -4% |
| Hispanic | *Within-ancestry* | 0.020 (0.001) | --- | -55% | -66% |
|  | *Cross-ancestry* | 0.046 (0.002) | 124% | --- | -24% |
|  | *Combined-ancestry* | 0.060 (0.003) | 197% | 32% | --- |
|  | *TL-GDES* | 0.058 (0.002) | 187% | 28% | -3% |
|  | *TL-PR* | 0.057 (0.002) | 181% | 25% | -5% |
|  | *TL-BMM* | 0.058 (0.002) | 186% | 28% | -4% |

*Within-ancestry* uses training data from AOU from the same ancestry as the testing data set. *Cross-ancestry* uses effects estimates estimated with data of EU-ancestry from the UK-Biobank. *Combined-ancestry* combines the data sets used in *within-ancestry* and *cross-ancestry* to estimate effects. The three Transfer Learning (TL-) methods use the estimates obtained with UK-Biobank data (*cross-ancestry*) as priors to a TL algorithm: Gradient Descent with Early Stopping, *GDES*, Penalized Regression, *PR*, or Bayesian Model, *BMM*, run on the data set used in the *within-ancestry* method.

**Figure 3.**
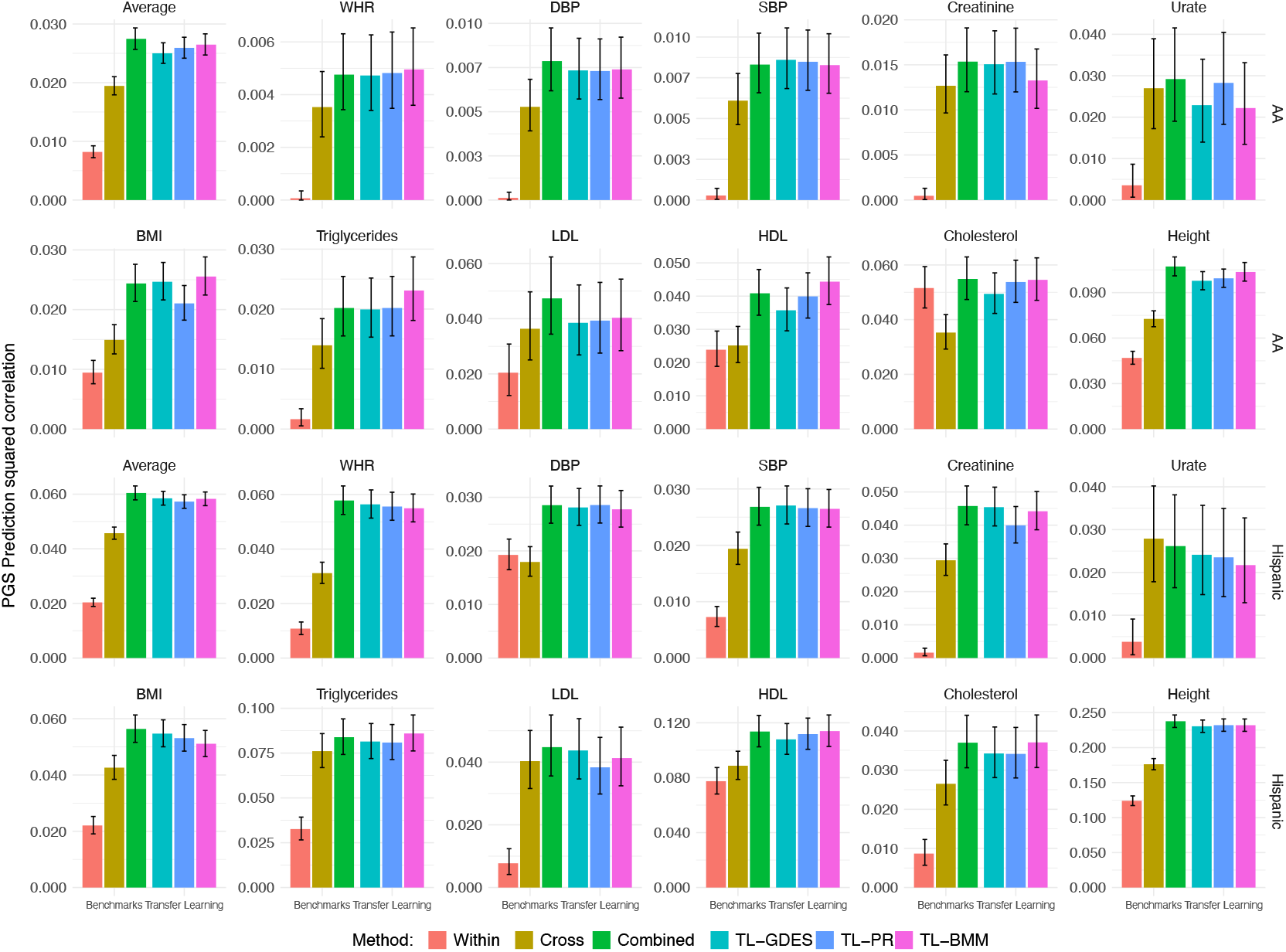
Prediction squared correlation (± SE) of PGS methods obtained with real phenotypes. *Within* uses training data from AOU from the same ancestry as the testing data set. *Cross* uses data of EU-ancestry from the UK-Biobank. *Combined* uses the data used in *Within* and *Cross*. The three Transfer Learning (TL-) methods use the estimates obtained with UK-Biobank data (*Cross*) as priors to a TL algorithm: Gradient Descent with Early Stopping, *GDES*, Penalized Regression, *PR*, or Bayesian Model, *BMM*, applied to the data set used in the *Within* method. WHR: waist-to-hip ratio, DBP: diastolic blood pressure, SBP: systolic blood pressure.

These results were based on averages across eleven traits; Figure 3 shows the results for each of the traits, target-ancestry groups, and methods. As expected, given the differences in heritability, the squared correlation varied substantially between traits; however, the ranking of the methods was similar across traits, with the *cross-ancestry* method outperforming the prediction performance of the *within*-*ancestry* PGS, the *combined-ancestry* PGS doing better than any of the two, and the TL methods performing similarly to the *combined-ancestry* prediction. The only traits that showed some (in many cases, small) departures from this trend were serum urate in Hispanics (where the *combined-ancestry* PGS and the TL methods were slightly worse than the *cross-ancestry* PGS), cholesterol in AA (for which the *within-ancestry* PGS performed better than the *cross-ancestry* PGS), and HDL and DBP in AA and Hispanics, respectively (for these traits, the *within-* and *cross-ancestry* PGSs achieved very similar prediction squared correlation).

#### Transfer Learning for EU target-ancestry using prior information derived from non-EU data

In the preceding analyses, we leveraged the large sample size available for EU ancestry to improve the prediction accuracy for non-EU groups using TL. We also explored whether TL from non-EU data can increase prediction accuracy for the EU group. To do this, we repeated analyses done to obtain the results in Figure 3, this time using the non-EU data (i.e., AA and Hispanic) as the source and EU (from the UK-Biobank) as the target populations. In this setting, we did not observe any improvements in PGS prediction accuracy when using TL relative to the within-ancestry prediction; this result is expected considering that the sample size for the target-ancestry is large and the sample size for the source-ancestry data was considerably smaller (**Figure S1**).

#### Sensitivity analyses with respect to the p-value threshold used to select variants

The results presented in Table 1 and Figure 3 used variants that had an association p-value < 1 × 10^−5^. We profiled these results, changing the p-value threshold from the GWAS-significant threshold (p-value < 5 × 10^−8^) to a very liberal threshold (p-value< 1 × 10^−3^). On average, increasing the p-value threshold above 1 × 10^−5^ did not result in sizable improvements in prediction accuracy in AA and led to a very small increase in prediction accuracy for Hispanics (**Figure S2**, Table S1). However, there were differences between traits worth mentioning. For highly complex traits such as height and BMI, using a more liberal p-value threshold led to increases in prediction accuracy; however, for lipids, increasing the p-value threshold above 1 × 10^−5^ resulted in a reduction in prediction accuracy.

Likewise, there were interesting differences between methods. Specifically, it seems that the combined analysis was more robust to differences in p-value threshold than the TL methods. For TL, using a moderate p-value threshold (e.g., 1 × 10^−5^) gives a good compromise of borrowing information without introducing in the PGS prior information about small effect variants, which may have poor portability between populations [18, 19].

#### Using sparse LD matrices may harm prediction accuracy in admixed populations

The computational burden of estimating effects grows exponentially with the number of variants used in the PGS. One way to reduce the computational cost of estimating hundreds of thousands of variant effects is to use sparse sufficient statistics or LD reference panels that account for local LD and ignore LD between distant variants. We compare the prediction accuracy of the PGS we developed using dense sufficient statistics with others derived using sparse sufficient statistics, where we zeroed out LD between chromosomes. For the *cross-ancestry* prediction (i.e., in the case where variant effects were estimated using data from EU-ancestry), we saw almost no difference in prediction accuracy between PGS using dense or sparse sufficient statistics. However, when the training of the PGS used data from non-EU ancestry (e.g., *within-, combined-ancestry* prediction, and all the TL methods), we observed a substantial reduction in PGS prediction accuracy when using sparse sufficient statistics. This reduction was particularly clear for Hispanics, an admixed population where LD between chromosomes may be present (**Figure S3**, Table S1).

#### Prediction performance of PGSs derived using data from multiple source-ancestry groups

The results presented before were based on PGS derived using data from one source-ancestry group. For the *combined-ancestry* and TL PGSs, we assessed whether training PGSs using data from multiple source-ancestry groups could improve prediction accuracy further. For these analyses we used dense LD matrices. On average, compared with the method using training data from single source-ancestry groups, combining data from multiple source-ancestry groups led to 21% (from 0.026 to 0.032) and 10% (from 0.058 to 0.064) gains in prediction squared correlations in AA and Hispanics, respectively (**Table 2**). These gains in prediction performance were consistent across traits and methods (**Figure 4**, Table S1).

**Table 2.** Average squared correlation (across 11 real phenotypes) by PGS methods.

| Target-Ancestry Group | Method | Average squared correlation (SE) |  | % Change |
| --- | --- | --- | --- | --- |
|  |  | Single source-ancestry group | Multiple source-ancestry groups |  |
| African American | *Combined-ancestry* | 0.027 (0.002) | 0.032 (0.002) | 16% |
|  | *TL-GDES* | 0.025 (0.002) | 0.031 (0.002) | 24% |
|  | *TL-PR* | 0.026 (0.002) | 0.032 (0.002) | 22% |
|  | *TL-BMM* | 0.026 (0.002) | 0.032 (0.002) | 21% |
| Hispanic | *Combined-ancestry* | 0.060 (0.003) | 0.066 (0.003) | 10% |
|  | *TL-GDES* | 0.058 (0.002) | 0.065 (0.003) | 11% |
|  | *TL-PR* | 0.057 (0.002) | 0.063 (0.003) | 10% |
|  | *TL-BMM* | 0.058 (0.002) | 0.063 (0.003) | 9% |

*Combined-ancestry* uses the data from UKB-EU, AOU-EU, AOU-AA, and AOU-Hispanics. The three Transfer Learning (TL-) methods use the estimates obtained from UKB-EU, AOU-EU, AOU-AA, and AOU-Hispanics as priors to a TL algorithm: Gradient Descent with Early Stopping, *GDES*, Penalized Regression, *PR*, or Bayesian Model, *BMM*.

**Figure 4.**
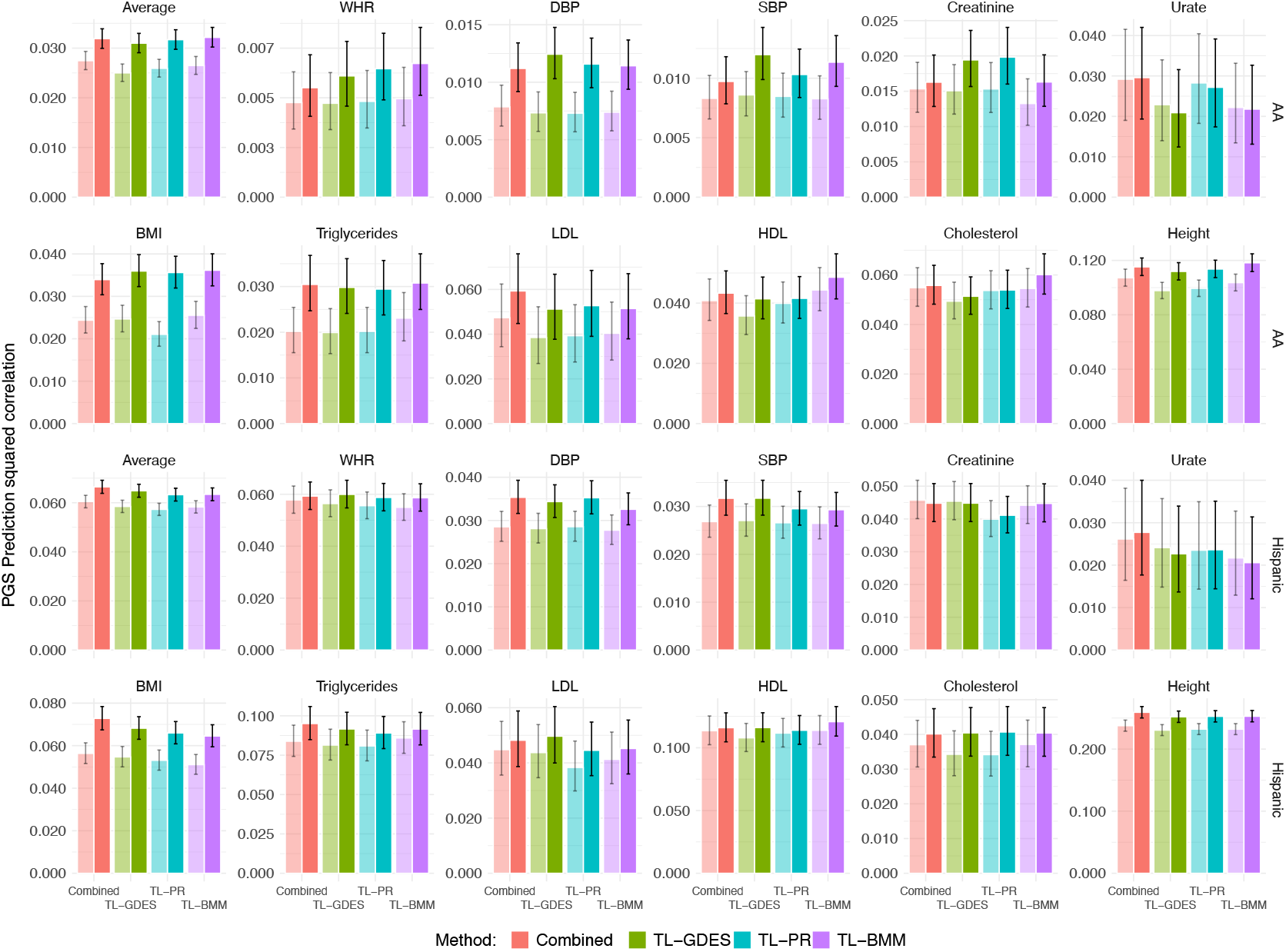
Prediction squared correlation (± SE) obtained when using data from a single source-ancestry group (left bars, light-shaded) versus using data from multiple source-ancestry groups (right bars, solid-shaded) by trait, method, and the ancestry of the testing data sets. For each of the methods used, the figure presents side-by-side bars representing the prediction Sq. Correlation (± SE) obtained when combining data from UK-Biobank plus data from the target-ancestry group (left light-shaded bars), and the one achieved when combining data (or summary statistics) from UK-Biobank, AOU-EU, and AOU target-ancestry group (right solid-shaded bars).

### Benchmark for PGS using all the available variants and sparse LD reference panels

Many investigations recommend building PGS using all the available variants (i.e., without performing variant selection using p-value thresholding) [2, 16, 17, 24]. In this case, because the number of variants is huge, sparse LD reference panels (accounting for within-block LD only) are typically used. Our previous results suggest that in the populations we are focusing on and with the sample sizes available, adding variants that have a GWAS p-value > 1 × 10^−5^ and using extremely sparse LD reference panels may reduce prediction accuracy for some traits. However, to demonstrate the feasibility of applying TL methods using nearly 1 million variants and to provide a benchmark of TL against PRS-CSx [16] and PROSPER [17]–two PGS methods for multi-ancestry data that use sparse LD reference panels–we present results from analyses done without applying p-value thresholding.

The Materials and Methods section titled Pipeline 2 provides details of how these analyses were done. Briefly, the inputs to these analyses were sparse LD reference panels for EU, AA, and Hispanics covering nearly 1 million variants and GWAS results obtained from the source population (EU UK-Biobank) and from training data in the target-ancestry groups (AA and Hispanics). These inputs were used to derive PGS using TL, PRS-CSx, and PROPSER, and the accuracy of these PGSs was evaluated in testing data from the target-ancestry groups.

PRS-CSx produces ancestry-specific estimates of effects; however, the developers of this method suggest building an ensemble of PGS from different ancestries; therefore, for PRS-CSx we present PGS prediction accuracies when using the target-ancestry PRS-CSx PGS as well as an ensemble of the PGSs from the source- and target-ancestry groups reported by PRS-CSx. To have a fair comparison, for TL we report results from the TL PGS and from an ensemble of that PGS and the source-population PGS. Finally, in the case of PROSPER, the ensemble step is part of the algorithm (referred to as “ensemble regression” in PROSPER); therefore, for this method we only present results for the ensemble PGS.

The results obtained using the analyses described above are presented in **Figure 5**. For both target-ancestries, on average, TL methods (especially *TL-BMM*) had higher prediction accuracy than PRS-CSx and PROSPER, and PRS-CSx slightly outperformed PROSPER (Figure 5, **Table S2**). The ranking of the three methods was rather consistent across traits and ancestry groups. The ensemble of EU and non-EU PGSs resulted in a substantial improvement in the prediction accuracy of the PGS derived using PRS-CSx; however, the ensemble of PGSs resulted in a very small further improvement in the accuracy of TL-PGS, suggesting that an ensemble step is not needed when TL methods are used.

Importantly, on average, the use of nearly 1 million variants and sparse LD reference panels in the PGS resulted in a reduction of prediction accuracy when compared with the average prediction performance of TL methods derived using variants with p-value < 1 × 10^−5^ and dense LD matrices (dashed horizontal lines in Figure 5, which correspond to the average results obtained with TL when using p-value thresholding and dense LD matrices, Figure 3).

**Figure 5:**
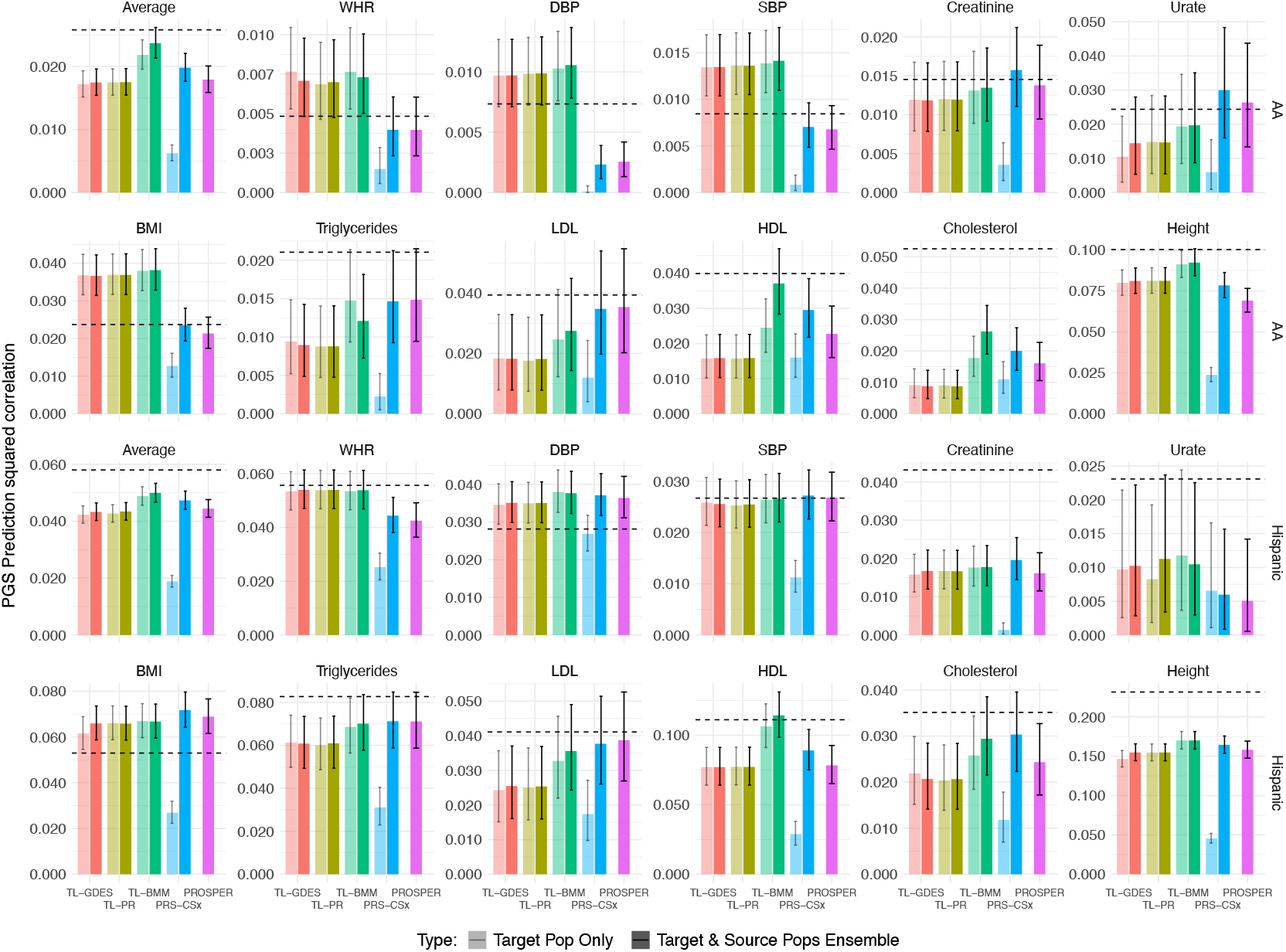
Prediction squared correlation (± SE) obtained when using all the available variants (minor-allele frequency > 0.01, ~1 million variants) and sparse LD reference panels, by statistical method, population, and trait. For each of the methods used, the figure presents side-by-side bars representing the prediction Sq. Correlation (± SE) obtained when using the estimated effects for the target population only (left light-shaded bars, except for PROSPER), and using an ensemble of the source- and target-ancestry PGSs (right solid-shaded bars). The horizontal dashed lines mark the averaged prediction Sq. Correlation obtained by using TL methods with p-value thresholding (GWAS-significant p-value < 1 × 1007) and dense sufficient statistics (Figure 3).

### Computational times

We performed a computational benchmark in Michigan State University’s high-performance computing cluster using nodes equipped with AMD EPYC 9654 Processor at 2.40 GHz and 768 GB of RAM. For a **first** set of **computational benchmarks**, we developed PGS varying the number of variants (**between 1**,**000 and 30**,**000 variants**) using **dense LD matrices**–this benchmark is relevant for situations where the PGS is derived using p-value thresholding and dense LD matrices are available. Within this setting, we recorded the time used by the GDES to complete 100 gradient descent cycles (usually early stopping happens before 10 or 20 cycles). For the PR we considered a grid of 100 values of *λ*, and for the BMM we simulated 5,000 posterior samples. All the computational times presented below are based on the average computing time over 10 independent runs of each of the algorithms.

Among the three TL algorithms, *TL-GDES* was the fastest; it takes less than a minute to complete 100 cycles of GD with tens of thousands of variants (**Table S3**). *TL-PR* is the most time-consuming of the three methods implemented in GPTL because many cycles (usually more than 300) of the coordinate descent algorithm are needed to achieve convergence for each value of λ. Thus, if we have 100 *λ* values and ~300 cycles are needed to converge, this method runs 30,000 times over each variant effect, while the GDES only passes 100 times for each effect. Finally, it took ~20 minutes for *TL-BMM* to simulate 5,000 posterior samples (Table S3). It is worth noting that the pipeline to implement *TL-BMM* is much simpler and requires a single run of the algorithm because regularization parameters and variant effects are jointly inferred from the posterior distribution. Thus, this method does not require calibrating regularization parameters.

The results presented above are all based on dense LD matrices. For any of the three methods, using **sparse LD matrices** can lead to remarkable reductions in the computational time required to fit a PGS. For instance, for the same number of variants (30,000), we observed a reduction in computing time greater than 95% when using a sparse LD matrix (with 95% of the cells being zero) compared with the time required when using a dense LD matrix (Table S3). However, as we noted earlier, using sparse LD matrices can also lead to a reduction in PGS prediction accuracy.

#### Computational benchmark of TL against PRS-CSx and PROSPER

For this comparison we recorded the computational times involved in deriving PGS in Pipeline 2 (nearly 1 million variants, 11 traits, two target-ancestry groups) for each of the methods (**Table S4**). Averaged across traits and target populations, the time used to run the TL algorithms ranged from 23 seconds (*TL-GDES*) to about 38 minutes (*TL-PR*). For the same task, it took much longer times to fit PGSs with PRS-CSx and PROSPER–about two orders of magnitude longer relative to *TL-PR* and *TL-BMM*, and three orders of magnitude longer than the time used by *TL-GDES*.

The computational time reported in Table S4 for TL methods takes into account the time needed to run the TL algorithm. If prior effect estimates are not available, there will be an additional computational time involved in deriving prior estimates of effects using summary statistics from the source populations. This additional computational time is the same as the one used by *TL-BMM* because one can derive those prior estimates by running *TL-BMM* with just one mixture component centered at zero. Even when considering this additional computational time, the TL algorithms implemented in GPTL are one or two orders of magnitude faster than PRS-CSx and PROSPER.

## Discussion

***Transfer Learning*** (TL) adapts knowledge gained using data from one population to improve the models’ performance on another population [25]. This approach is commonly used in machine learning and can be used to improve PGS prediction accuracy for groups that are underrepresented in GWAS data sets.

We present GPTL, an R-package including three methods to develop PGS using TL. The main functions, GD(), PR(), and BMM() take sufficient statistics (SS, ***X′ X, X′y***) as inputs. The SS can be derived from individual genotype data or from LD reference panels (dense or sparse) and GWAS results. Any of the three methods can be used to fit PGS with large numbers of variants (millions of variants if the LD-reference panel is sparse) and biobank-scale data.

The three TL algorithms that we implemented in the GPTL R-package induce shrinkage of variant effect estimates toward prior values through different mechanisms. In ***TL-GDES*** [9], shrinkage is controlled by the number of iterations run before early stopping and the learning rate. In general, as one would expect, in cases where the *cross-ancestry* PGS performed well, the algorithm stopped at early stages; thus, inducing strong shrinkage towards initial values (**Figure S4**).

In ***TL-PR***, shrinkage is achieved by penalizing deviations of estimates relative to prior values. Here, we also observed that in cases where the *cross-ancestry* PGS performed well, the optimal *λ*-value was high; thus, inducing strong shrinkage towards the prior values (**Figure S5**). Furthermore, for most trait-ancestry combinations, the optimal *α*-values were 0 (Ridge penalty), indicating that the models were shrinking towards prior values without setting any estimate exactly equal to the EU-derived effects used as prior values (this could only happen when *α* > 0). The only exceptions were total cholesterol and HDL cholesterol in AA, where the optimal *α*-values were equal to 1 (Lasso penalty, compare height and total cholesterol in **Figure S6** for contrasting examples).

PROSPER also uses penalized regression to derive PGS from multi-ancestry data. The objective function used in PROSPER penalizes both effect sizes (i.e., the distance between estimates and null effects) and differences of effects between populations. Thus, PROSPER shrinks the effects of both the source and target populations towards common values and both towards zero. Our *TL-PR* is slightly different in that TL uses estimates derived from a source population (e.g., EU) as prior values to an algorithm that runs on data from the target population.

Finally, in ***TL-BMM***, shrinkage towards prior values is controlled by the mixture prior used, which has one component with zero mean (this component induces shrinkage towards null effects), and another component centered at the EU-derived variant effect estimates. In this case, as one would expect, the posterior probability of the component centered at EU-derived variant effects was higher for ancestry-traits combinations where the *cross-ancestry* PGS had a relatively good performance (**Figure S7**).

Our empirical results show that the extent to which prior effects are shrunk towards prior values varies between traits, algorithms, source- and target-ancestry groups, and the sample size of the target ancestry. For instance, TL estimated effects were closer to prior values when the **target-ancestry** was Hispanic than AA (compare, for the same trait, the results for AA and Hispanics displayed in Figure S6), suggesting that, as one would expect, EU effects are more portable to Hispanics than to AA.

Likewise, we observed smaller changes in effects (from prior values to TL estimates) for traits with a small training **sample size** (e.g., cholesterol) than for more heritable traits with a larger sample size (e.g., height, see Figure S6). A large sample size and a highly heritable trait resulted in a very sharp objective function (or posterior distribution in the case of BMM), which enables stronger departures of TL estimates from prior values.

The **comparison between methods** suggested that, for any trait-population combination, the BMM was the method producing stronger differences between prior values and TL estimates of effects. This is likely due to the use of a two-component finite mixture prior, which allows BMM to explore models ranging from the cross-ancestry (this will happen when the component centered at prior values dominates) and the within-ancestry (which will happen when the component with zero prior mean dominates).

Finally, we also noticed that the **larger-effect variants are more likely to persist**, and the smaller-effect variants tend to have larger differences between prior values and TL estimates of effects (Figure S6). This result is in agreement with previous studies suggesting that the portability of effects between populations is better for large-effect loci than for those with small effects [18, 19].

### Transfer Learning achieves results similar to those of a combined analysis without requiring the sharing of individual data or large sufficient statistics

Our analyses showed that *combined-ancestry* PGS outperformed *within-* and *cross-ancestry* PGSs [23], and the three TL methods achieved a prediction accuracy competitive with that of the *combined-ancestry* PGS. Importantly, the TL methods only require sharing variant effect estimates, while implementing the *combined-ancestry* PGS can be challenging and may pose privacy concerns because it requires sharing individual genotype and phenotype data or large sufficient statistics.

While on average, TL methods performed similarly to PGS derived using *combined-ancestry* data, there were interesting differences in the relative performance of these methods between traits and populations. These differences are likely due to the underlying assumptions of these methods. Specifically, a *combined-ancestry* PGS assumes that effects are the same in all populations; however, TL allows for effect heterogeneity. Therefore, a *combined-ancestry* PGS works best when effects are very similar between populations. Thus, in our simulation, when the effect heterogeneity at causal variants was low (i.e., causal effects correlation = 0.95), *combined-ancestry* PGS was slightly better than TL PGS, and when effect heterogeneity was substantial (i.e., causal effects correlation = 0.6), TL PGS achieved a much higher prediction accuracy than *combined-ancestry* PGS. Interestingly, even in scenarios where effect-heterogeneity was very low (i.e., causal effects correlation = 0.95), for African Americans, we observed that TL methods outperformed the *combined-ancestry* PGS; this did not happen in Hispanics [19]. This is because, although causal loci effects were almost the same in the two populations, owing to differences in LD patterns and allele frequencies, marker effects can still differ [18, 19].

Consistent with the results of our simulations, in real data, we observed that the *cross-ancestry* PGS performed relatively well in Hispanics and less well in AA. Therefore, the gains in prediction accuracy brought by TL methods were relatively higher in AA compared to Hispanics. However, there were important differences between traits. For instance, for human height, a trait that previous studies suggest has similar variant effects across populations [18, 19], we observed that *combined-ancestry* PGS achieved a higher prediction accuracy than TL PGSs. Conversely, for lipid traits, TL PGS performed better than the *combined-ancestry* PGSs in AA. Previous studies reported substantial variant effect-heterogeneity between AA and EUs for lipid traits [18]. TL seems a good strategy for these traits because it allows borrowing information (shrinking estimates towards prior values) without assuming that the effects are the same across populations.

### Using all the available variants is not always best

Many previous studies suggest that PGS using all variants achieves higher prediction accuracy than those derived using p-value thresholding [2, 16, 24, 26]. However, most of these studies are based on analysis of highly complex traits (such as height or BMI) using biobank-size data from EU ancestry. Our study suggests that when deriving TL-PGSs for non-EU populations (particularly Hispanics), a strategy that uses all the available variants and sparse LD reference panels leads, on average, to lower prediction accuracy than the one that can be achieved using fewer variants (selected through p-value thresholding) and dense LD matrices (compare the dashed horizontal lines and the bars in Figure 5). This observation can be attributed to multiple factors. Firstly, while ignoring LD between blocks may not be a big problem in EU populations, not accounting for LD between blocks seems to have a sizable negative effect on prediction accuracy in an admixed population such as Hispanics (Figure S3, Table S1). Secondly, previous studies suggest that large effects are more portable between populations than small effects [18, 19]. Therefore, it seems reasonable to expect that including large numbers of very small-effect variants (which happens when p-value thresholding is not used) can reduce the accuracy of a PGS derived through TL. Thirdly, when the sample size of the target-ancestry group is small (as was the case for lipid traits in our study), using all the available variants may result in low prediction accuracy because when the number of variants is too large relative to the sample size, variant effects are estimated with very low accuracy [27, 28].

### Internal vs. external LD reference panels

The software we provide can be used with both internal and external reference panels. In the analyses we present in this study, we used internal LD reference panels (i.e., LD matrices derived with the same data used to obtain GWAS results) because a large fraction of the variants available in the target data (AOU) were not present in pre-existing LD reference panels. In general, one would expect results to be slightly worse when external reference panels are used if the sample size used to derive some LD reference panels is small (e.g., 1000 Genomes [29]) and in cases where there are mismatches between the ancestry of the data used to obtain the GWAS results and the one used to derive the LD reference panel.

### Transfer Learning performs an automatic ensemble

Many PGS methods [9, 16, 17, 30–34], including PRS-CSx and PROSPER, utilize an ensemble process to improve the prediction accuracy by averaging PGSs derived from both the source- and target-ancestry groups. The TL methods presented in our study achieve such an ensemble process automatically by shrinking the estimated effects for the target population towards the values obtained in the source population. Therefore, in our benchmarks with PRS-CSx, the ensemble process brought large improvements in the prediction accuracy for ancestry-specific PGS derived using PRS-CSx but only slight increases for TL methods (Figure 5). Moreover, the coefficients of the OLS ensembles (Table S2) had values near zero for the EU PGS when the PGS was from a TL algorithm, and large values when the PGS being averaged was derived using PRS-CSx.

### Transfer Learning between genetically correlated traits

In this study, we focused on TL (for the same trait) between populations. Another possible use of TL would be to transfer knowledge between genetically correlated traits. For instance, one can imagine having variant effect estimates from PGS published for traits that are genetically correlated to Type 2 Diabetes (T2D, e.g., BMI, blood glucose, and potentially many traits related to metabolic syndrome) as prior information to a TL implemented to develop a PGS for T2D. For such applications, *TL-BMM* appears as a potentially attractive model given its ability to incorporate prior information from multiple sources. In general, such applications of TL would be more promising when the sample size for the target trait or disease (e.g., T2D) is smaller than the sample size available for the traits whose estimates are used as prior information.

### Decentralized and Federated Learning

Several studies (e.g., [9, 23]), including ours, show that combining information from multiple data sets leads to sizable improvements in PGS prediction accuracy. As more data sets are created (and grown) in the private and public domains, the opportunities to further improve PGSs by using data from multiple sources will continue to increase. However, analyzing these datasets jointly will become increasingly difficult and, in many instances, impossible due to privacy concerns. Transfer Learning algorithms can be an effective tool for implementing Federated Learning, a model used in some machine learning applications that enables collaborative model training across distributed datasets without requiring direct data sharing [35].

## Materials and Methods

### Datasets

In this study, we used data from the UK-Biobank and the All of Us (AOU) cohorts.

#### Sample selection in the UK-Biobank

From this cohort, we used data from distantly related Europeans *n*~312,000. We excluded samples of individuals who withdrew from the study and those who did not have data for any of the eleven phenotypes used in analyses.

#### Sample selection in All of Us

We included in the data analyses *n*~79,000 European Americans, *n*~30,000 African Americans, and *n*~28,000 Hispanics from the AOU cohorts (CDRv7, Controlled Tier). The sample size (number of subjects with phenotype and genotype data) varied between traits (**Table S5**). Within African American and Hispanic groups, individuals were randomly assigned to either a training set (80%) or a testing set (20%).

#### Genotypes

For analysis involving the variants from the AOU cohort, we had 1,675,354 variants from the AOU arrays in autosomal chromosomes. For the UK-Biobank cohort, we used 771,437 variants in autosomal chromosomes that were present (matched using rs-IDs) in the AOU array and in the imputed UK-Biobank genotypes. We checked for the consistency of the genotyped strand and the reference alleles; when opposite reference alleles were used in UK-Biobank and AOU, variant effects estimates derived using UK-Biobank data were multiplied by −1.

#### Phenotypes

For the real-data analyses, we used phenotypes that were available in the UK-Biobank and AOU cohorts. These included three anthropometric traits (height, BMI, waist-to-hip ratio), six metabolic biomarkers (serum urate, creatinine, triglycerides, and total, HDL, and LDL cholesterol), and two vital measurements (systolic and diastolic blood pressures).

#### Phenotype extraction, transformations, and units

For the UK-Biobank, we extracted phenotypes from visit one or (if there was no data at visit one) visit two. For AOU, when multiple records were available, we used the one that was closest to the genotyping date. For blood pressure measurements, we used weekly averages and extracted the data from the week that was closest to the genotyping date. When the units of the phenotype were different in UK-Biobank and AOU, we converted the phenotypes to the units used in the UK-Biobank. **Table S6** provides the units for each of the traits analyzed. Serum urate and creatinine were highly skewed; therefore, these two phenotypes were log-transformed.

### Pipeline 1: benchmark using variants selected through p-value thresholding

The pipeline described in Figure 1 was used to benchmark PGS methods using variants selected through GWAS analysis with p-value thresholding. We benchmarked methods using simulated and real phenotypes. Unless it is otherwise indicated, in Pipeline 1, we used dense LD-matrices derived from either UK-Biobank or AOU data.

#### Simulation of phenotypes using All of Us genotypes

For simulations, we generated quantitative phenotypes using genotypes in the All of Us (AOU) cohort from 151,025 European American (EA), 57,331 African American (AA), and 54,640 Hispanic subjects. Using the genotypes of these individuals (*p*=1,675,354 variants), we simulated a trait with 500 causal variants and a trait heritability of 0.4. We excluded from the simulation variants with minor-allele frequency smaller than 0.5% and those with more than 3% of missing values.

Rather than simulating a trait using the entire genome, for each of the ten Monte Carlo replicates, we simulated and analyzed data by chromosomes. This was done in the following steps:

1. Within each chromosome *k* (*k* = 1, …, 22), we sampled at random *q*_*k*_ = ⌊500(*p_k_*/*p*) variants, where ⌊⋅⌋ denotes the floor function, *p_k_* was the number of variants in the chromosome *k*, and *p* was the total number of variants available.
2. We sampled allele effects for the causal variants in the source-(*s*, EA) and target-ancestry (*t*, AA or Hispanic) groups from IID bivariate normal distributions with zero mean, variances equal to one, and effect correlations (ρ) equal to either 0.95 or 0.6, that is

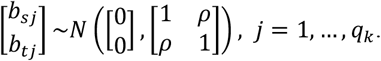
3. We then formed the additive genetic values in the source-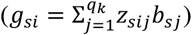 and target-ancestry 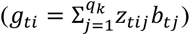 populations as linear combinations of the genotypes at the causal variant (z_∗*ij*_).
4. Finally, we formed phenotypes by adding to the genetic values IID normal error terms with error variances calibrated to match a chromosome heritability of 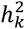 where 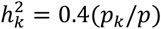.

The analysis pipeline for the simulated phenotypes was the same as the one used for real phenotypes (more details are provided below). Importantly, we excluded from the analyses (both in the GWAS and effect estimation steps) the genotypes at the causal variants.

The prediction accuracy of the resulting PGS was evaluated using a testing set consisting of 10,000 randomly sampled subjects whose data were not used in any of the steps involved in the derivation of the PGSs. The prediction accuracy reported was the squared correlation between the PGS and the simulated genetic values, divided by the chromosome heritability 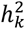

#### Phenotype pre-processing in real-data analyses

##### UK-Biobank

For this data set, each phenotype was pre-adjusted using an ordinary least squares regression including sex, age, the first five variant-derived principal components, center, and batch. Since we used only data from individuals of European ancestry, the PCs here account for the sub-structure within that ancestry group.

##### All of Us

Unlike the UK-Biobank, in AOU, there were a few clear outliers which were likely the result of recording errors; therefore, for this data set, we remove phenotypes that deviated from the median more than five interquartile range (i.e., the difference between the 75^th^ and the 25^th^ percentiles). Subsequently, we pre-adjusted each of the eleven phenotypes using an ordinary least squares regression including sex, age, and self-reported ancestry (which matched very closely the clustering based on variant-derived PCs).

#### Analysis details for Pipeline 1

The training data sets consisted of individuals of European ancestry (in simulations: EAs from AOU; in real data analyses: EUs from the UK-Biobank) and 80% of the participants of non-EU target-ancestry from AOU (Hispanic or AA). We used the training data for variant selection and effects estimation. We evaluated the prediction performance of the resulting PGS in the remaining 20% of the non-EU target-ancestry data in AA and Hispanics from AOU that was not used for model training.

Our analysis **Pipeline 1** consists of three steps (Figure 1). In **Step 1**, we selected the variants to be included in the PGS based on GWA analyses done using data from the source-ancestry (EU) and the training data from the target-ancestry group (either AA or Hispanic). We used in further analyses variants with GWAS p-value < 1 × 10^−5^ (in a sensitivity analysis we profiled the value of the p-value threshold from the standard GWAS-significance threshold, p-value < 5 × 10^−8^ to a very liberal threshold, p-value < 1 × 10^−3^). The number of variants for different methods and p-value thresholds is listed in **Table S1**. In **Step 2**, we estimated effects using various strategies: for the *cross-ancestry* and *within-ancestry* PGSs we estimated effects by fitting a Bayesian regression [36, 37] to the training data sets from the source population and target populations, respectively. For the *combined-ancestry* PGS, the same model was applied to a data set that combined the training data sets from the source and target populations. For the TL methods we used the estimated effects in constructing *cross-ancestry* PGS as prior information to a TL algorithm applied to the training data set from the target population. Finally, in **Step 3**, we evaluated the PGS prediction accuracy in the testing data set of the target-ancestry–these data were not used in Steps 1 and 2. Next, we provide additional information about each of the steps.

##### Step 1: GWAS and variant selection

For each phenotype, we first conducted separate GWA analyses in the UK-Biobank (EU ancestry), AOU European American data, AOU African American and Hispanic training data sets, separately. For each GWAS, the association between pre-adjusted phenotypes and variants was assessed using a (one-variant-at-a-time) least squares regression. These analyses were done using the BGData [38] R-package.

The variants used to build PGS were those with a GWAS p-value < 1 × 10^−5^ (or < {5 × 10^−8^, 1 × 10^−5^, 5 × 10^−5^, 1 × 10^−4^, 5 × 10^−4^, 1 × 10^−3^} in an analysis that we did for the real phenotypes to assess the sensitivity of results with respect to the p-value threshold used). The GWAS results used to perform variant selection varied between methods (**Figure 5**).

**Figure 5:**
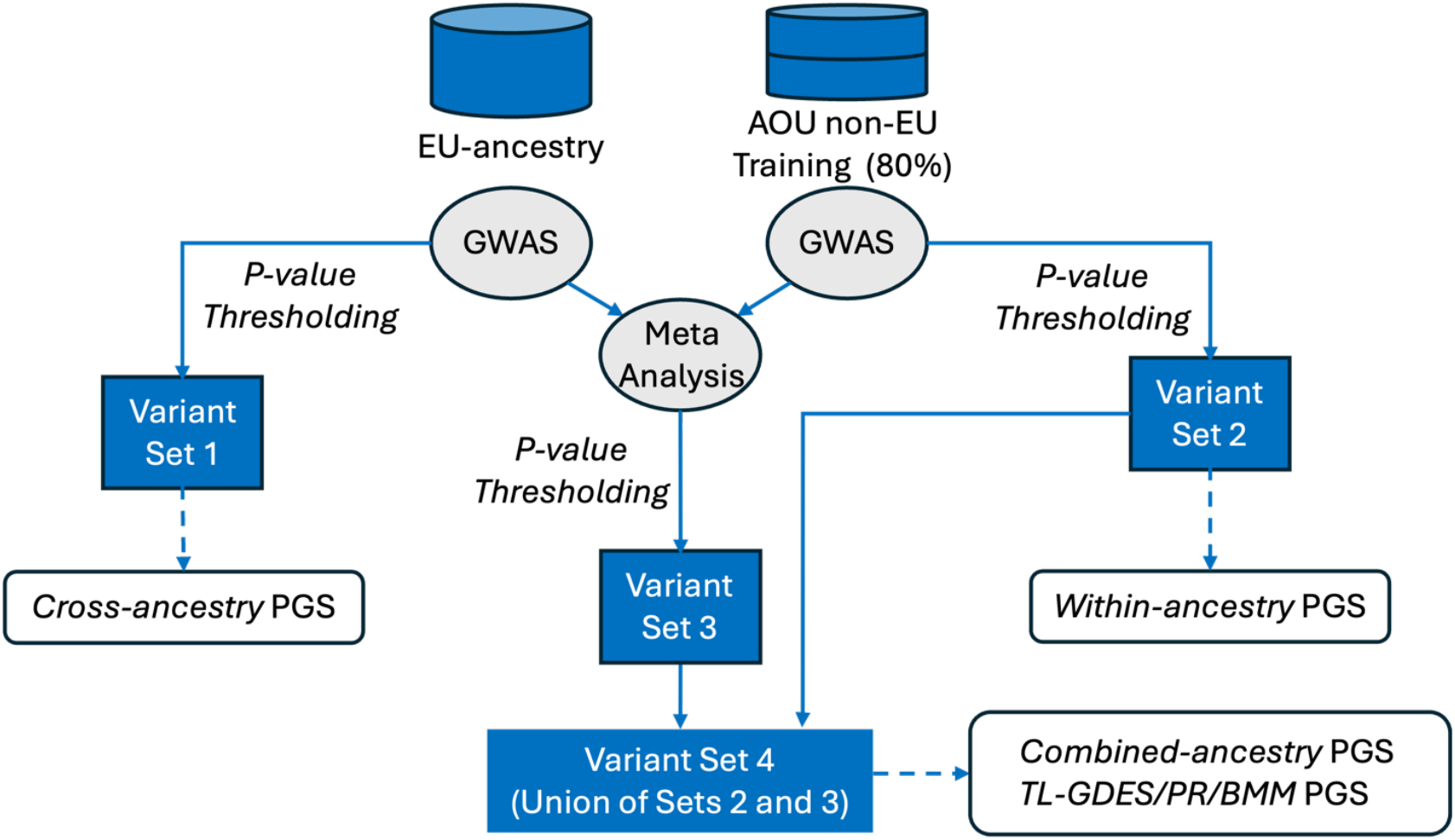
Graphical representation of variant selection (Pipeline 1). Sets of variants were selected from different GWAS results. To derive *cross-* and *within-ancestry* PGSs, variants were directly selected by p-value threshold from the GWASs EU (Set 1) and AOU non-EU training (Set 2) data sets, respectively. To derive *combined-ancestry* and TL-based PGSs, a meta-analysis first selected variants with a significant pooled effect (Set 3). Then the variants in the union of Sets 2 and 3 will be used for constructing PGS.

For *cross-ancestry* PGS prediction, the variants were selected using UK-Biobank EU-ancestry GWAS results. For *within-ancestry* PGS prediction, variant selection was based on the GWAS done in the training data from the target-ancestry group. Finally, for TL PGS and *combined-ancestry* PGS, we meta-analyzed the GWAS results from EU and non-EU ancestry. The meta-analysis t-statistics (H_0_: T_*j*_ = 0) for the variant *j* were

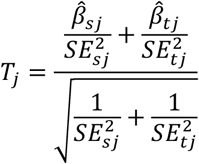

where 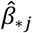 and 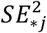 are the estimated effect and the SE for the variant *j* in either the source-(*=s) or target-ancestry (*=t) population. Subsequently, a two-sided p-value was obtained from a standard normal distribution.

#### Variant selection when using data from multiple source-ancestry groups

Above, we described the procedure used for variant selection for PGS that involved data from one source-ancestry group. We also evaluated the *combined*-*ancestry* or TL PGS derived using data from multiple source-ancestry groups. In this case, we first meta-analyzed the GWAS results of all the ancestry groups involved (using a procedure like the one described above), and the variant set used was the union of the variants with significant association (p-value < 1 × 10^−5^) in either the meta-analysis or the GWAS done in the training data of the target-ancestry group

##### Step 2: Variant effects estimation

For the selected significant variants (variant Set 4), we derived sufficient statistics from the training data sets (SS, ***X***′***X*** and ***X***′***y***, separate SS were derived for each ancestry group using the same samples as in the GWAS), and used these SS to estimate variant effects. For all the analyses presented in Pipeline 1 (except those in Figure S3), the sufficient statistics consisted of dense cross products of centered genotypes (***X***′***X***, which, up to scale, is a dense LD matrix), and cross products between genotypes and adjusted phenotypes (***X***′***y***). For the results presented in Figure S3 and in Tables S1 (analyses labeled as sparse by chromosome) and S3 (analyses indicated as sparse), we zero out LD between chromosomes, thus making ***X***′***X*** block-diagonal with one non-zero block per chromosome.

For the *cross-ancestry* PGS we estimated effects using EU-ancestry derived SS (variant Set 1, UK-Biobank in the real data analysis and European Americans from AOU in the simulations), for *within-ancestry* we used the AOU SS of the corresponding ancestry group (variant Set 2, either AA or Hispanic), and finally, for the *combined ancestry* we estimated effects by combining (adding) SS used in *cross-* and *within-ancestry* PGSs. For these three benchmarks (*combined*-, *within-*, and *cross-ancestry* PGS), we estimated effects using a Bayesian shrinkage estimation method (a Bayesian model with a Gaussian prior centered at zero, model ‘BRR’ in the BGLR R-package [36, 37]). For the TL algorithms, we used the EU-derived estimates (i.e., the ones used in *cross-ancestry* PGS) as prior for a TL algorithm that iterated on training data of non-EU ancestry.

#### Calibration of shrinkage parameters

The *TL-GDES* and *TL-PR* require tuning the shrinkage parameters (the learning rate and the number of iterations in *TL-GDES*, and the regularization and penalty parameters in *TL-PR*, respectively). To do this, we did a 2-fold cross-validation within the training data set. Then, for each fold, we fitted models over a grid of values of the shrinkage parameters using one fold and evaluated prediction accuracy in the other fold. We repeated this by switching the fold assignments. Using the results from this calibration analysis, we chose the optimal values of the shrinkage parameters and fitted a model to the entire training data set using the chosen shrinkage parameter values.

In ***TL-GDES***, to select the optimal number of iterations, in each run, one calibration set was first used to train the model for a user-defined number of iterations (by default, 100 iterations). The 100 sets of estimated effects were used to predict the phenotype in the second calibration set and evaluate the prediction accuracy. The 2-fold cross-validation will generate two sets of 100 prediction accuracy curves, which will be averaged to select the optimal number of iterations that achieves the largest prediction accuracy.

In ***TL-PR***, two shrinkage parameters (*α* and *λ*) need to be selected. For the *α*, we used a grid of equally distributed values between 0 (i.e., ridge regression penalty) and 1 (i.e., lasso regression penalty): {0, 0.2, 0.4, 0.6, 0.8, 1}. For Lasso and Elastic net models (*α* ≠ 0), following Friedman, Hastie, and Tibshirani’s idea [11], we used the following strategy to generate a grid of 100 *λ*-values. The steps are described using R’s programming language syntax.

If *α* ≠ 0,

> lambda.max=(max(abs(Xy))+1e-5)/alpha
>
> lambda.min=lambda.max/100
>
> lambda.grid=exp(seq(from=log(lambda.max),to=log(lambda.min),length=100))

For Ridge Regression models, we first constructed a grid of R-sq. values (from 0.01 to 0.9, equally spaced in the log scale) and then derived a grid of values for *λ*.

If *α* = 0,

> grid=exp(seq(from=log(10000),to=log(1),length=100))
>
> K=mean(diag(XX))
>
> lambda.grid=K*grid

#### Variant effects estimation when using data from multiple source-ancestry groups

For the *combined-ancestry* PGS using data from multiple source-ancestry groups, we estimated effects by adding the SS from the four groups mentioned above. For instance, to predict Hispanics, we combined the SS of EU (UK-Biobank), EA, AA, and 80% of the Hispanic data from AOU. Likewise, to estimate the effects using the *combined-ancestry* method for AA, we obtained the SS by combining SS derived from EU (UK-Biobank), EA, Hispanics, and 80% of the AA data from AOU.

For TL methods using data from multiple source-ancestry groups, we derived the prior effects estimates using data from three groups. For example, for the prediction of Hispanics, we derived separate variant effects estimates using data from EU (UK-Biobank), EA, and AA from AOU. Then, for *TL-GDES* and *TL-PR*, the prior information used was an average of these three estimates. For *TL-BMM*, we used a mixture prior with four components, three using the prior means derived from EU, EA, and AA data, and a fourth one having a null mean. We ran the TL algorithms on 80% of the Hispanic data from AOU, and prediction accuracy was evaluated in the 20% Hispanic data that was left out. The scheme for AA was similar; however, in this case we obtained the prior information from EU (UK-Biobank), EA, and Hispanic data from AOU, we ran the TL algorithms on 80% of the AA data, and we assessed prediction accuracy in the remaining 20% of AA data from AOU.

##### Step 3: Evaluation of prediction performance

We evaluated the PGS prediction accuracy in the testing data (either AA or Hispanic from AOU) that was not used in any of the previous steps of the pipeline (either GWAS, variant selection, or effects estimation). This was done by correlating PGS predictions with adjusted phenotypes. We evaluated the prediction performance via the squared correlation.

### Pipeline 2: benchmark using all the available variants without applying p-value thresholding

We also benchmarked TL-PGS constructed using all the available variants (i.e., without p-value thresholding), and compared prediction performances among TL methods, PRS-CSx, and PROSPER. The pipeline used in this case was very similar to the one described above with benchmarks using p-value thresholding (Pipeline 1, Figure 1); however, in Pipeline 2 we: (i) use all the variants that were available in AOU (after standard QC and minor-allele frequency filtering, more below), (ii) variant effects were estimated using sparse LD reference panels (that account for LD within blocks and ignore LD between blocks) and GWAS summary statistics, (iii) in the case of BMM, for reasons that we explain below, the BMM variance components were profiled, and (iv) in addition to target-ancestry PGSs, we also considered using an ensemble of EU and target-ancestry PGS–a strategy recommended for PRS-CSx and automatically generated by PROSPER.

#### Phenotype-preprocessing

was the same as described for Pipeline 1.

#### Variant QC

For AOU target-ancestry groups, we included variants with a missing frequency < 0.03 and minor-allele frequency > 0.01. This resulted in 993,783 variants for AA and 875,221 variants for Hispanics. For the UK-Biobank, we included 771,437 variants that are present in the above AOU variant lists and in the imputed UK-Biobank genotypes.

#### Training, calibration, and testing sets

To train models we used the same training set as the one used in Pipeline 1. However, for this pipeline, to calibrate shrinkage and ensemble parameters, we further split the testing set used in Pipeline 1 (20% of the target-ancestry data from AOU) into equally large calibration and testing sets, each having ~10% of the available data for the ancestry group in AOU. We used the calibration set to profile shrinkage parameters (e.g., early stopping, regularization parameter in the TL-PR, and variance components for BMM, more on this below) as well as the weights used for ensemble methods.

#### LD-reference panels

PRS-CSx and PROSPER provide LD reference panels for several ancestry groups constructed using UK Biobank data [16]. However, only ~250,000 of the variants in these reference panels were available in AOU, and the ancestries with LD reference panels in PRS-CSx and PROSPER (EU, African, American, and Asian) do not match the target-ancestry groups of our study (AA and Hispanics) well. Therefore, we constructed new reference panels for AA and Hispanics using AOU data, maintaining the LD-block definition (the start and end base pair position of each of the blocks) of the panels provided by PRS-CSx. The resulting AOU-derived LD reference panels include 5,080 blocks for AA and 5,055 blocks for Hispanic ancestries. These LD reference panels are provided with this manuscript (see Data and code availability). For TL we used these reference panels in the TL step, and for PRS-CSx and PROSPER, we used these reference panels together with the EU reference panel provided by PRS-CSx

##### Step 1: GWAS

The methodology was as described in Step 1 of Pipeline 1. Here, instead of using the results to select variants (p-value thresholding), we used the GWAS summary statistics as input to the algorithms (along with sparse LD reference panels) to estimate variant effects

##### Step 2: Variant effects estimation and model parameter calibration

#### Prior effects for TL methods

We first estimated the EU-ancestry effects using UK-Biobank data. The estimation method and software were the same as the ones used for prior effects in Pipeline 1; however, here, considering the large number of effects that needed to be estimated jointly (~900,000), we fitted models within overlapping chromosome segments using the approach used in [39] and [40]. Briefly, models were fitted to chunks of 5,000 variants; this window was displaced by 1,000 variants. From each segment we retrieved effect estimates from the core of 3,000 variants and discarded estimates from the flanking regions (1,000 variants in each flanking region). We refer to [39] and [40] for further details. This procedure provided prior estimates which were used in TL methods. For the variants not present in UK-Biobank, we used a prior value of 0.

#### TL-PGS effects

These were estimated using the prior effect estimates, the target-ancestry sparse LD reference panels, and GWAS results from the training data set of the target-ancestry group. To produce the inputs for the GPTL functions, we used the function getSS() of the GPTL R-package. This function takes as inputs the GWAS results and the LD reference panel, and produces the inputs (a sparse ***X***′***X*** and a dense ***X***′***y***) used in GD(), PR(), and BMM().

For *GDES*, we recorded the estimated effects for each of the first 50 cycles of the GD algorithm. We then profiled prediction accuracy in the calibration set by cycle and chose an optimal early stopping.

For *PR*, we set *α* = 0 (Ridge regressions) and fitted models over a grid of 30 *λ*-values. We then profiled prediction accuracy in the calibration set versus *λ* to choose an optimal value from where we obtained the effects for the *TL-PR*.

For *BMM*, we made slight modifications compared to Pipeline 1 to accommodate the use of sparse sufficient statistics. The unknown parameters of the BMM model include the intercept, marker effects, mixture probabilities, and variance parameters. To update the error variance, during MCMC cycles, *BMM* internally updates the residual sum of squares using sufficient statistics

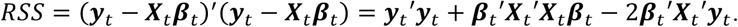

When ***X***_*t*_′***X***_*t*_ is a dense matrix, RSS can be computed properly. However, when ***X***_*t*_′***X***_*t*_ is a sparse matrix that ignores the between-block LD, the equality (***y***_*t*_ − ***X***_*t*_*β*_*t*_)′(***y***_*t*_ − ***X***_*t*_*β*_*t*_) = ***y***_*t*_′***y***_*t*_ + *β*_*t*_′***X***_*t*_′***X***_*t*_*β*_*t*_ − 2*β*_*t*_′***X***_*t*_′***y***_*t*_ does not hold. Thus, when sparse SS are used, variance parameters cannot be properly updated. To circumvent this problem, we fixed the error and effects variances using 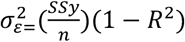 and 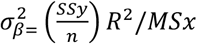 respectively, where *R*^2^ is the proportion of the phenotypic variance explained by the model, *SSy* is the sum of squares of the phenotype (after pre-adjusting by covariates), *n* is the training sample size, and *MSx* is the sum of the variances of the variants included in the model. The values of the variance parameters depend on the unknown *R*^2^; therefore, we ran *BMM* with fixed variance parameters over a grid of values of *R*^2^ ∈ {0.002, 0.005, 0.01, 0.02, 0.05, 0.1,0.2,0.5}, evaluated prediction accuracy on the calibration set, and selected effects from the run with the optimal *R*^2^ for predictions for the *TL-BMM* model.

#### PRS-CSx effects

We used the GWAS results from AOU training data and UK-Biobank, together with the corresponding LD reference panels as inputs of PRS-CSx to estimate effects. We used the default parameter settings as recommended by PRS-CSx (n_iter=1,000, n_burnin=500, thin=5). PRS-CSx provides estimates of variant effects for each of the ancestry groups (EU and the target ancestry, either AA or Hispanics).

#### Ensemble of ancestry-specific PGS for PRS-CSx and TL

PRS-CSx produces estimates of effects for the EU and non-EU ancestry groups. Likewise, for TL we have EU effects (used as prior means) and target-ancestry effects derived using the TL algorithms, which run on target-ancestry training data. Following the recommendations made by the developers of PRS-CSx [16], we built the final ensemble as a weighted sum of the PRS-CSx EU and target-ancestry PGSs with weights equal to the OLS coefficients estimated in a calibration data set. To have a fair comparison, we also built a similar ensemble PGS for the TL methods using the UK-Biobank PGS and the TL PGS.

#### Ensemble of ancestry-specific PGS for PROSPER

In this case, the ensemble is part of the PROPSER algorithm/pipeline; therefore, we simply used the estimated effects reported by PROPSER after the super-learning step to compute PGS for PROSPER.

##### Step 3: Evaluation of prediction performance

We evaluated the PGS (both the target-ancestry PGSs and the ensemble PGSs) prediction accuracy in testing data from AOU that was not used in any steps involved in deriving PGS (GWAS, profiling of key parameters, or the ensemble of PGS). We evaluated the prediction performance via the squared correlation between PGS predictions and adjusted phenotypes.

## Data Availability

All data produced in the present work are contained in the manuscript.

https://doi.org/10.5281/zenodo.16923734

https://doi.org/10.5281/zenodo.17087604

## Data and code availability

The UK-Biobank and All of Us cohort data sets are available under restricted access; access can be obtained by applying at https://www.ukbiobank.ac.uk/ and https://allofus.nih.gov. The raw UK-Biobank and the All of Us cohort data are protected and are not available due to data privacy laws.

The LD reference panels generated during this study are available at https://doi.org/10.5281/zenodo.16923734, and the GWAS results are available at https://doi.org/10.5281/zenodo.17087604.

The software presented and described in this study is available as an R-package (GPTL), which can be downloaded and installed from https://github.com/QuantGen/GPTL/. The script generated during this study is available at https://doi.org/10.5281/zenodo.17177288.

## Acknowledgments

We thank the participants of the UK-Biobank and All of Us studies and the scientists and staff involved in participant recruitment and data collection. H.W. and G.D.L.C. received funding from grants NSF 2035472 and USDA/NIFA 2023-70412-41054. H.W., A.I.V., and G.D.L.C. received funding from grant HG013794 and from Michigan State University. M.B. received funding from grant HG009976.

## Author contributions

Conceptualization: H.W. and G.D.L.C.; Methodology: H.W., M.B., and G.D.L.C.; Software Development: H.W., P.P.R, and G.D.L.C.; Investigation: H.W. and G.D.L.C.; Writing-Original Draft: H.W.; Writing-Review & Editing: H.W., P.P.R, M.B., Y.C., X.L., A.I.V., and G.D.L.C.; Project Administration and Supervision: G.D.L.C.; Funding Acquisition: G.D.L.C. All authors read and approved the final manuscript.

## Competing interests

The authors declare no competing interests.

## Notes

### Competing Interest Statement

The authors have declared no competing interest.

### Funding Statement

This study was funded by grants NSF 2035472 and USDA/NIFA 2023-70412-41054.

### Author Declarations

The study used UK-Biobank and All of Us cohort data sets. The UK-Biobank and All of Us cohort data sets are available under restricted access; access can be obtained by applying at https://www.ukbiobank.ac.uk/ and https://allofus.nih.gov.

