## Supplementary Figures for "Improving Polygenic Score Prediction for Underrepresented Groups Through Transfer Learning"

**Supplemental Figures**

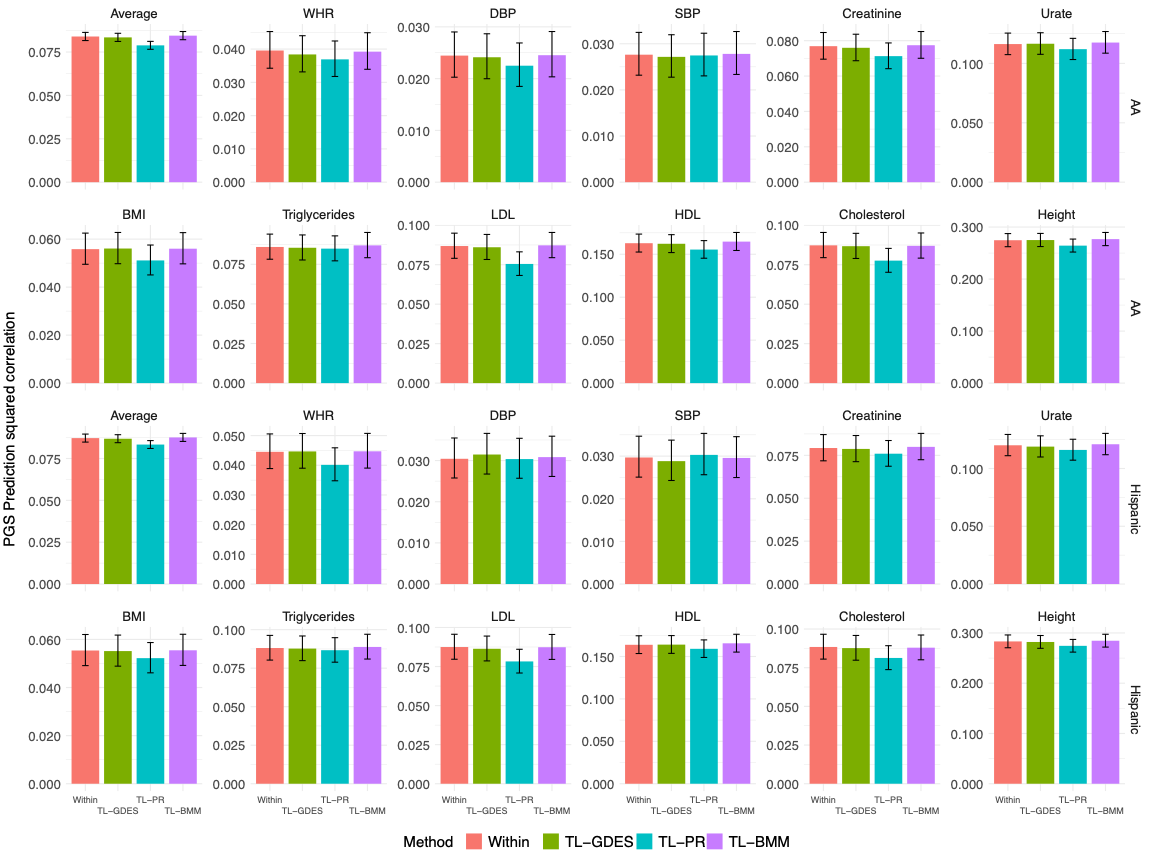

**Figure S1. Prediction squared correlation (± SE) obtained when taking AA/Hispanic as the source population and EU as the target population.** P-value thresholding (p-value $<1\times{10}^{-5}$) was used to select variants. Within uses training data from the UK Biobank (EU) as the testing data set. The three Transfer Learning (TL-) methods use the estimates obtained with AOU data (AA or Hispanic) as priors to a TL algorithm: Gradient Descent with Early Stopping, GDES, Penalized Regression, PR, or Bayesian Model, BMM, applied to the data set used in the Within method.

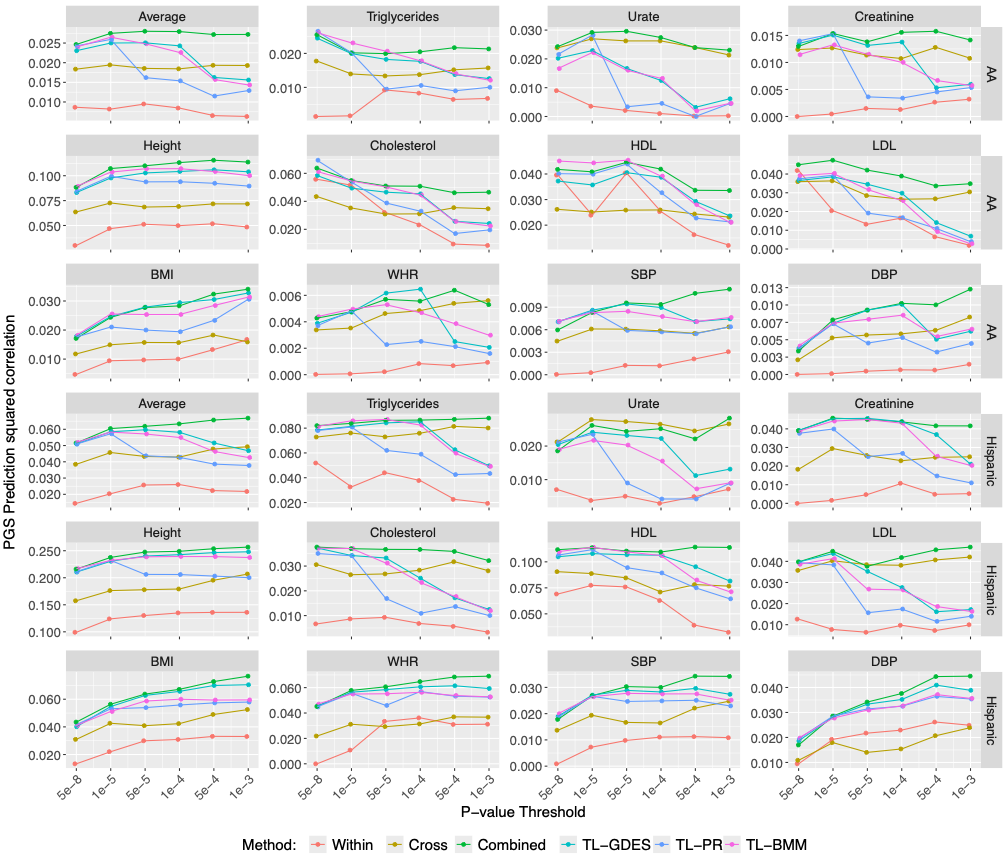

**Figure S2. Prediction squared correlation obtained when using variants selected by different p-value thresholds, varied by trait, method, and the ancestry of the testing data sets.** For each of the methods used, the figure presents the prediction Sq. Correlation obtained when using variants selected by p-value $<\{5\times{10}^{-8},{1\times10}^{-5},{5\times10}^{-5},{1\times10}^{-4},{5\times10}^{-4},{1\times10}^{-3}\}$. Within uses training data from AOU from the same ancestry as the testing data set. Cross uses data of EU-ancestry from the UK-Biobank. Combined uses the data used in Within and Cross. The three Transfer Learning (TL-) methods use the estimates obtained with UK-Biobank data (Cross) as priors to a TL algorithm: Gradient Descent with Early Stopping, GDES, Penalized Regression, PR, or Bayesian Model, BMM, applied to the data set used in the Within method.

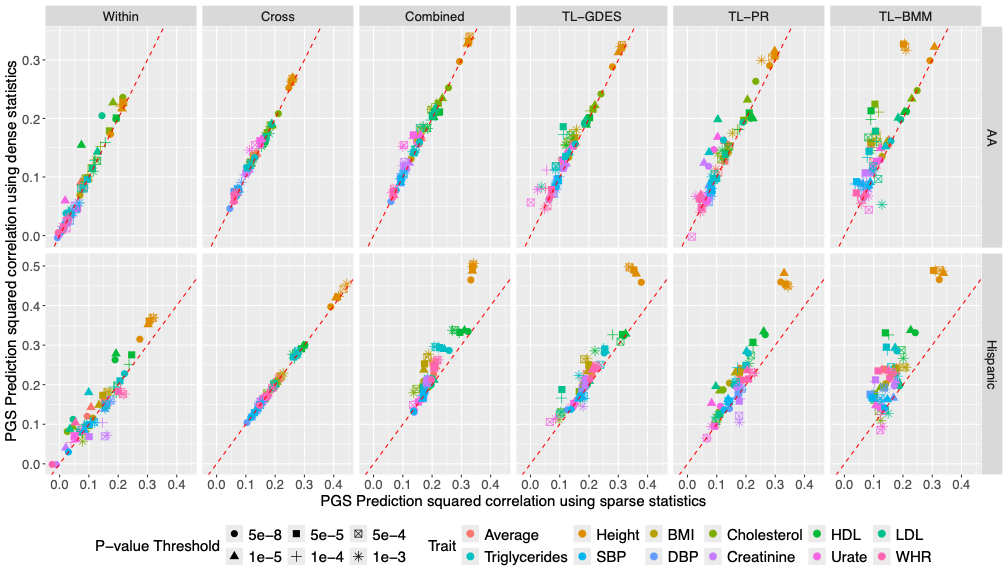

**Figure S3. Prediction squared correlation obtained when using dense or sparse statistics, varied by p-value threshold, trait, method, and the ancestry of the testing data sets.** For each of the methods used, the figure presents the prediction Sq. Correlation obtained when using sparse statistics (x-axis) or dense statistics (y-axis). Each dot corresponds to a trait, and each dot shape corresponds to a p-value threshold $\{5\times{10}^{-8},{1\times10}^{-5},{5\times10}^{-5},{1\times10}^{-4},{5\times10}^{-4},{1\times10}^{-3}\}$ for variant selection. Within uses training data from AOU from the same ancestry as the testing data set. Cross uses data of EU-ancestry from the UK-Biobank. Combined uses the data used in Within and Cross. The three Transfer Learning (TL-) methods use the estimates obtained with UK-Biobank data (Cross) as priors to a TL algorithm: Gradient Descent with Early Stopping, GDES, Penalized Regression, I, or Bayesian Model, BMM, applied to the data set used in the Within method.

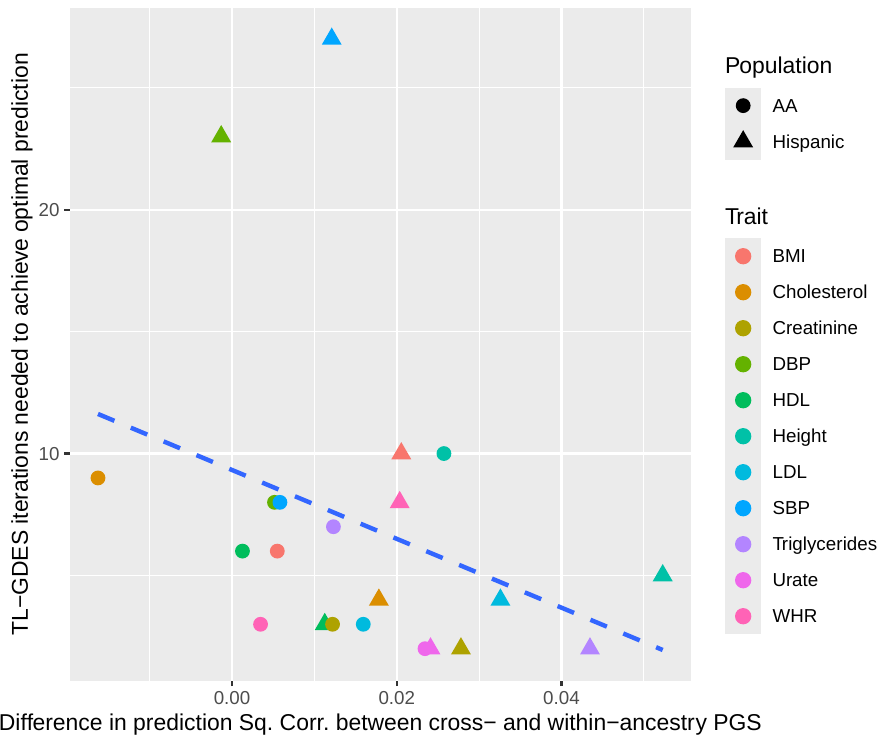

**Figure S4. The number of iterations required to achieve optimal prediction accuracy in TL-GDES.** Each dot corresponds to a trait-ancestry combination. The dashed line represents the ordinary least squares fit.

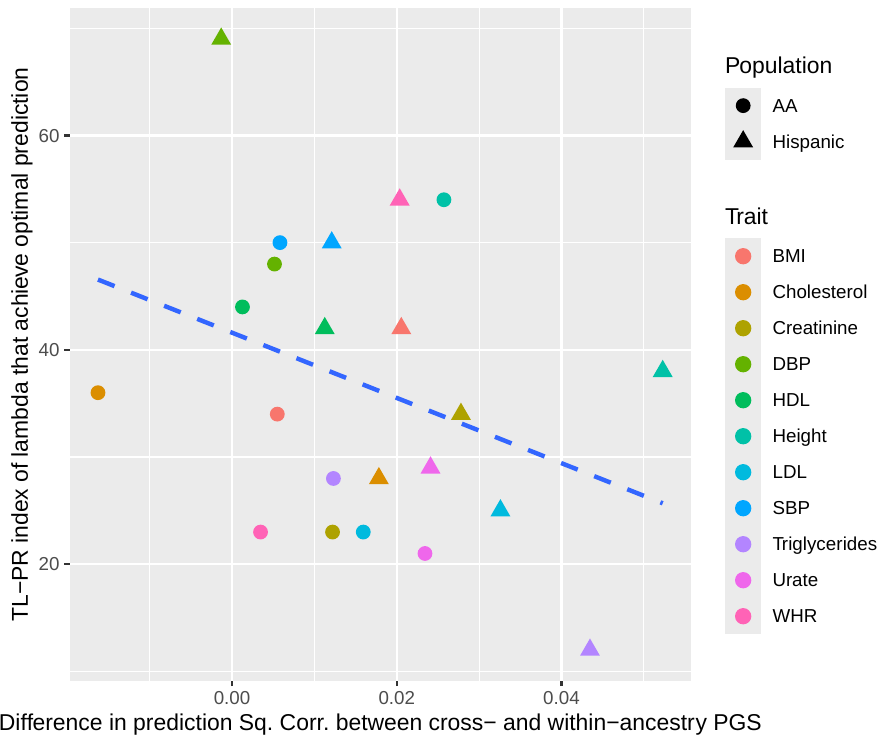

**Figure S5. The index of the optimal** $\boldsymbol{\lambda}$ **parameter in TL-PR models.** A larger index corresponds to a smaller $\lambda$ value. Each dot corresponds to a trait-ancestry combination. The dashed line represents the ordinary least squares fit.

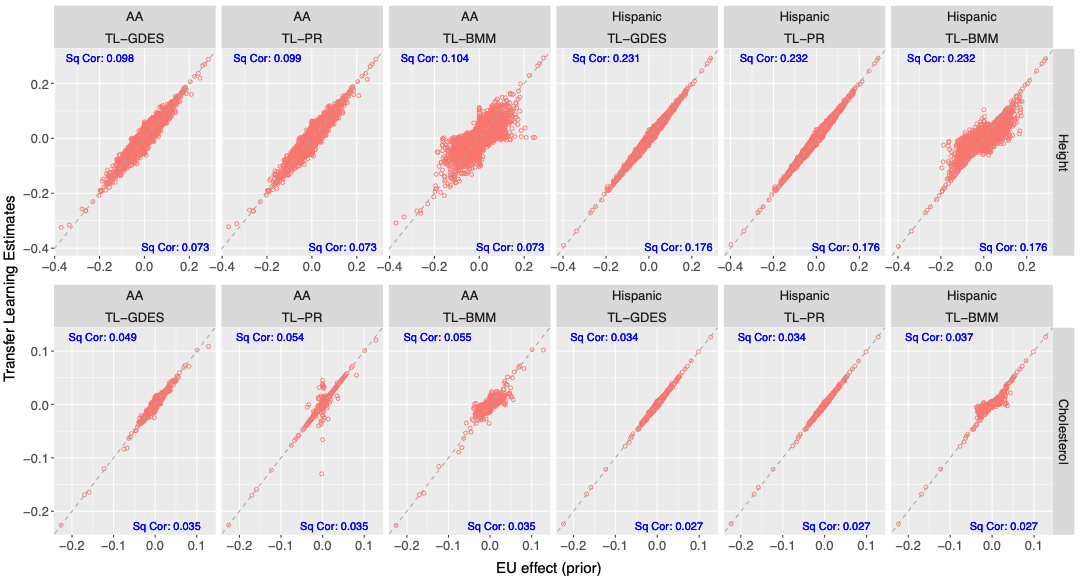

**Figure S6. Estimated variant effects using TL methods versus the European-derived estimates used as prior values, varied by trait, method, and the ancestry of the testing data sets.** Each circle corresponds to a variant in the PGS. Prediction Squared Correlations from the European-derived estimates are marked in the lower right corner, while Prediction Squared Correlations from the TL methods-derived estimates are marked in the upper left corner.

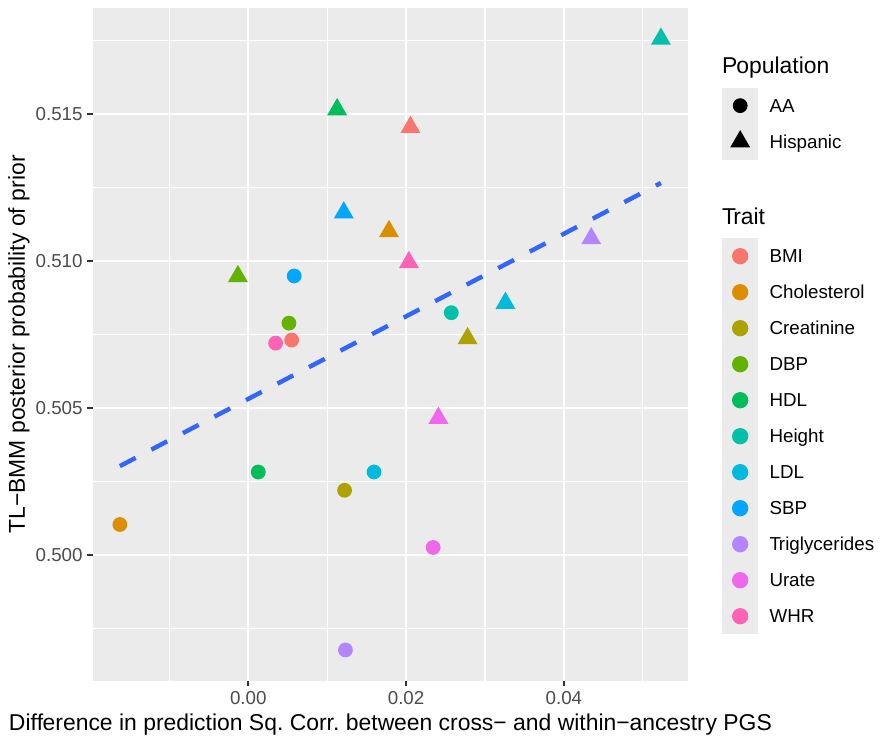

**Figure S7. The posterior probability of inclusion of prior in TL-BMM models.** Each dot corresponds to a trait-ancestry combination. The dashed line represents the ordinary least squares fit.
