## Supplementary Methods for "Improving Polygenic Score Prediction for Underrepresented Groups Through Transfer Learning"

**Coordinate Descent algorithm for the informed penalized regression**

In this section, we present the formulae used to implement the coordinate descent algorithm for the penalized regression. Recall that the loss function to be minimized, i.e., equation (1) in the manuscript is

$$\begin{aligned} L\left( \boldsymbol{\beta}_{t} \right)={\frac{1}{2}\left( \boldsymbol{y}_{t}-\boldsymbol{X}_{t}\boldsymbol{\beta}_{t} \right)}^{'}\left( \boldsymbol{y}_{t}-\boldsymbol{X}_{t}\boldsymbol{\beta}_{t} \right)+\alpha\lambda\parallel\boldsymbol{\beta}_{t}-{\hat{\boldsymbol{\beta}}}_{s}\parallel_{1}+\frac{1}{2}\left( 1-\alpha\right)\lambda\left( \boldsymbol{\beta}_{t}-{\hat{\boldsymbol{\beta}}}_{s} \right)^{'}\left( \boldsymbol{\beta}_{t}-{\hat{\boldsymbol{\beta}}}_{s} \right), \end{aligned}$$

where $\boldsymbol{y}_{t}$ and $\boldsymbol{X}_{t}$ are the vector of phenotypes and the genotype matrix for the training set of the target-ancestry population (e.g., Hispanics), respectively, $\boldsymbol{\beta}_{t}$ is the (unknown) vector of variant effects in the target-ancestry population, ${\hat{\boldsymbol{\beta}}}_{s}$ is the vector of prior variant effect estimates (e.g., variant effects derived using data from European ancestry), and $\alpha\in[0,1]$ and $\lambda>0$are shrinkage parameters.

After expanding the quadratic forms in (1) and removing the terms that do not involve the unknown coefficients ($\boldsymbol{\beta}_{t}$), the objective function can be written as

$L\left( \boldsymbol{\beta}_{t} \right)\propto\frac{1}{2}\left( \boldsymbol{\beta}_{t}'\boldsymbol{C}\boldsymbol{\beta}_{t}-2\boldsymbol{\beta}_{t}'\boldsymbol{r} \right)+\lambda_{1}\left| \boldsymbol{\beta}_{t}-{\hat{\boldsymbol{\beta}}}_{s} \right|+\frac{1}{2}\lambda_{2}\boldsymbol{\beta}_{t}'\boldsymbol{\beta}_{t}-\lambda_{2}\boldsymbol{\beta}_{t}'{\hat{\boldsymbol{\beta}}}_{s}$,

where $\boldsymbol{C}=\boldsymbol{X}_{t}'\boldsymbol{X}_{t}$ and $\boldsymbol{r}=\boldsymbol{X}_{t}'\boldsymbol{y}_{t}$ , $\lambda_{1}=\alpha\lambda$, and $\lambda_{2}=\left( 1-\alpha\right)\lambda$.

The partial derivative of $L\left( \boldsymbol{\beta}_{t} \right)$ function w.r.t to $\boldsymbol{\beta}_{t}$ is

$\frac{\partial L\left( \boldsymbol{\beta}_{t} \right)}{\partial\boldsymbol{\beta}_{t}}=\boldsymbol{C}\boldsymbol{\beta}_{t}-\boldsymbol{r}+\lambda_{1}\frac{\partial\left| \boldsymbol{\beta}_{t}-{\hat{\boldsymbol{\beta}}}_{s} \right|}{\partial\boldsymbol{\beta}_{t}}+\lambda_{2}\boldsymbol{\beta}_{t}-\lambda_{2}{\hat{\boldsymbol{\beta}}}_{s}$.

Therefore, the partial derivative w.r.t. to the $j^{th}$ coefficient is

$\frac{\partial L\left( \boldsymbol{\beta}_{t} \right)}{\partial\beta_{tj}}=C_{jj}\beta_{tj}+\Sigma_{k\neq j}C_{jk}\beta_{tk}-r_{j}+\lambda_{1}\frac{\partial\left| \beta_{tj}-\hat{\beta}_{sj} \right|}{\partial\beta_{tj}}+\lambda_{2}\beta_{tj}-\lambda_{2}\hat{\beta}_{sj}$.

After grouping terms, we get

$$\begin{aligned} \frac{\partial L\left( \beta_{t} \right)}{\partial\beta_{tj}}=\left( C_{jj}+\lambda_{2} \right)\beta_{tj}-\left( \tilde{r}_{j}+\lambda_{2}\hat{\beta}_{sj} \right)+\lambda_{1}\frac{\partial\left| \beta_{tj}-\hat{\beta}_{sj} \right|}{\partial\beta_{tj}},\#(S1) \end{aligned}$$

where $\tilde{r}_{j}=r_{j}-\Sigma_{k\neq j}C_{jk}\beta_{tk}$.

To obtain $\frac{\partial\left| \beta_{tj}-\hat{\beta}_{sj} \right|}{\partial\beta_{tj}}$ we must consider three cases:

**Case 1**

If $\beta_{tj}<\hat{\beta}_{sj}$, then $\frac{\partial\left| \beta_{tj}-\hat{\beta}_{sj} \right|}{\partial\beta_{tj}}=-1$; using this in equation (S1) we get

$\frac{\partial L\left( \beta_{t} \right)}{\partial\beta_{tj}}=\left( C_{jj}+\lambda_{2} \right)\beta_{tj}-\left( \tilde{r}_{j}+\lambda_{1}+\lambda_{2}\hat{\beta}_{sj} \right)$.

Setting the derivative equal to zero and solving for $\beta_{tj}$ we get

Update Case 1:

$$\begin{aligned} \beta_{tj}=\frac{\tilde{r}_{j}+\lambda_{2}\hat{\beta}_{sj}+\lambda_{1}}{C_{jj}+\lambda_{2}}.\#(S2) \end{aligned}$$

Using the above equation in the condition $\beta_{tj}<\hat{\beta}_{sj}$ we get

$$\frac{\tilde{r}_{j}+\lambda_{2}\hat{\beta}_{sj}+\lambda_{1}}{C_{jj}+\lambda_{2}}<\hat{\beta}_{sj}\Leftrightarrow\tilde{r}_{j}+\lambda_{2}\hat{\beta}_{sj}+\lambda_{1}<C_{jj}\hat{\beta}_{sj}+\lambda_{2}\hat{\beta}_{sj}.$$

Subtracting $\lambda_{2}\hat{\beta}_{sj}$ from both sides of the inequality and subsequently dividing both sides by $C_{jj}$ we get

$\beta_{OLS_{j}}+\frac{\lambda_{1}}{C_{jj}}<\hat{\beta}_{sj}$,

where $\beta_{OLS_{j}}=\tilde{r}_{j}/C_{jj}$. Therefore, the condition for case 1 can be expressed as

Condition Case 1:

$$\begin{aligned} \beta_{OLS_{j}}-\hat{\beta}_{sj}<\frac{\lambda_{1}}{C_{jj}}.\#(S3) \end{aligned}$$

**Case 2**

The derivative of the absolute value function is undefined at 0 (which, in our case, happens when $\beta_{tj}=\hat{\beta}_{sj}$). Therefore, we must consider the set of gradients that occur at that point and check whether the set includes a stationary point.

At $\beta_{tj}=\hat{\beta}_{sj}$, the set of gradients is $\frac{\partial\left| \beta_{tj}-\hat{\beta}_{sj} \right|}{\partial\beta_{tj}}\in\left[ -1,1 \right]$. Using this in (S1) gives the following set of gradients

$\frac{\partial L\left( \boldsymbol{\beta}_{t} \right)}{\partial\beta_{tj}}\in\left[ \left( C_{jj}+\lambda_{2} \right)\beta_{tj}-\left( \tilde{r}_{j}+\lambda_{1}+\lambda_{2}\hat{\beta}_{sj} \right), \left( C_{jj}+\lambda_{2} \right)\beta_{tj}-\left( \tilde{r}_{j}-\lambda_{1}+\lambda_{2}\hat{\beta}_{sj} \right) \right]$.

For a stationary point to exist within this set the following conditions must be met

$\left( C_{jj}+\lambda_{2} \right)\beta_{tj}-\left( \tilde{r}_{j}+\lambda_{1}+\lambda_{2}\hat{\beta}_{sj} \right)\leq0\leq\left( C_{jj}+\lambda_{2} \right)\beta_{tj}-\left( \tilde{r}_{j}-\lambda_{1}+\lambda_{2}\hat{\beta}_{sj} \right)$.

Using $\beta_{tj}=\hat{\beta}_{sj}$

$$\left( C_{jj}+\lambda_{2} \right)\hat{\beta}_{sj}-\left( \tilde{r}_{j}+\lambda_{1}+\lambda_{2}\hat{\beta}_{sj} \right)\leq0\leq\left( C_{jj}+\lambda_{2} \right)\hat{\beta}_{sj}-\left( \tilde{r}_{j}-\lambda_{1}+\lambda_{2}\hat{\beta}_{sj} \right).$$

Therefore,

$C_{jj}\hat{\beta}_{sj}-\tilde{r}_{j}-\lambda_{1}\leq0\leq C_{jj}\hat{\beta}_{sj}-\tilde{r}_{j}+\lambda_{1}$.

Dividing by $C_{jj}$ we get

$$\hat{\beta}_{sj}-\beta_{OLS_{j}}-\frac{\lambda_{1}}{C_{jj}}\leq0\leq\hat{\beta}_{sj}-\beta_{OLS_{j}}+\frac{\lambda_{1}}{C_{jj}}.$$

Adding $\beta_{OLS_{j}}-\hat{\beta}_{sj}$ we get

$$-\frac{\lambda_{1}}{C_{jj}}\leq\beta_{OLS_{j}}-\hat{\beta}_{sj}\leq\frac{\lambda_{1}}{C_{jj}}.$$

Therefore,

Condition Case 2:

$$\begin{aligned} \left| \beta_{OLS_{j}}-\hat{\beta}_{sj} \right|\leq\frac{\lambda_{1}}{C_{jj}}.\#(S4) \end{aligned}$$

In which case we will have

Update Case 2:

$$\begin{aligned} \beta_{tj}=\hat{\beta}_{sj}.\#(S5) \end{aligned}$$

**Case 3**

If $\beta_{tj}>\hat{\beta}_{sj}$, then $\frac{\partial\left| \beta_{tj}-\hat{\beta}_{sj} \right|}{\partial\beta_{tj}}=1$. Using this in equation (S1) we get $\frac{\partial L\left( \boldsymbol{\beta}_{t} \right)}{{\partial\beta}_{tj}}=\left( C_{jj}+\lambda_{2} \right)\beta_{tj}-\left( \tilde{r}_{j}-\lambda_{1}+\lambda_{2}\hat{\beta}_{sj} \right)$.

Setting the derivative equal to zero and solving for $\beta_{tj}$ we get

Update Case 3:

$$\begin{aligned} \beta_{tj}=\frac{\tilde{r}_{j}+\lambda_{2}\hat{\beta}_{sj}-\lambda_{1}}{C_{jj}+\lambda_{2}}.\#(S6) \end{aligned}$$

Using the above equation in condition $\beta_{tj}<\hat{\beta}_{sj}$ we get

$\frac{\tilde{r}_{j}+\lambda_{2}\hat{\beta}_{sj}-\lambda_{1}}{C_{jj}+\lambda_{2}}<\hat{\beta}_{sj}\Leftrightarrow\tilde{r}_{j}+\lambda_{2}\hat{\beta}_{sj}-\lambda_{1}<C_{jj}\hat{\beta}_{sj}+\lambda_{2}\hat{\beta}_{sj}$.

Subtracting $\lambda_{2}\hat{\beta}_{sj}$ from both sides of the inequality and subsequently dividing both sides by $C_{jj}$ we get $\beta_{OLS_{j}}-\frac{\lambda_{1}}{C_{jj}}<\hat{\beta}_{sj}$. Therefore,

Condition Case 3:

$$\begin{aligned} \beta_{OLS_{j}}-\hat{\beta}_{sj}<-\frac{\lambda_{1}}{C_{jj}}.\#(S7) \end{aligned}$$

Therefore, the conditions and the updates needed to implement a coordinate descent algorithm to minimize the objective function (1) can be summarized as follows.

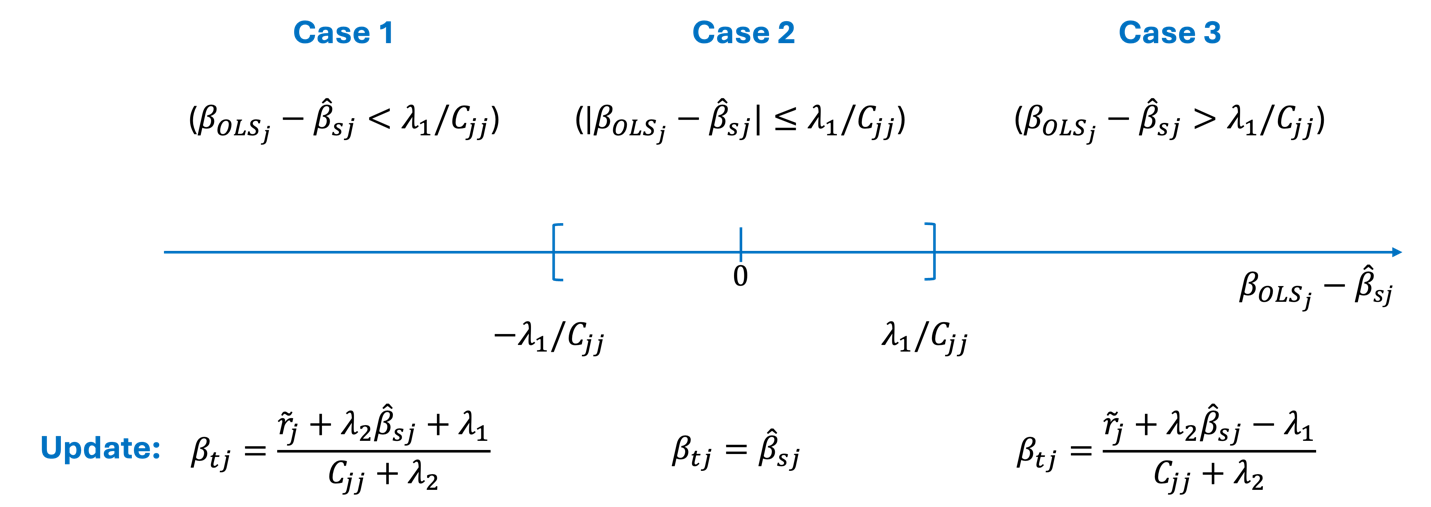

**Graphical representation of the Coordinate Descent algorithm.** The three cases are depicted above the axis and the three types of updates are below the axis. The horizontal axis represents $\beta_{OLS_{j}}-\hat{\beta}_{sj}$.

**Coordinate Descent algorithm updates**

Using the conditions (expressions (S3), (S4), and (S7)) and updates described above (equations (S2), (S5), and (S6)), we have a Coordinate Descent algorithm that can be implemented using the following updates of individual coefficients.

- Compute $\beta_{OLS_{j}}=\frac{r_{j}-\Sigma_{k\neq j}C_{jk}\beta_{jk}}{C_{jj}}$,
- If $\left| \beta_{OLS_{j}}-\hat{\beta}_{sj} \right|<\frac{\lambda_{1}}{C_{jj}}$,

set $\beta_{tj}=\hat{\beta}_{sj}$,

Else

set $\beta_{tj}=\frac{\tilde{r}_{j}+\lambda_{2}\hat{\beta}_{sj}-sign\left( \beta_{OLS}-\hat{\beta}_{sj} \right)\lambda_{1}}{C_{jj}+\lambda_{2}}$.
